# Repeated Handgrip Strength Variability in Myalgic Encephalomyelitis/Chronic Fatigue Syndrome: Separating Disease-Related Fatigability from Force Gradation

**DOI:** 10.64898/2026.07.22.26358672

**Authors:** Felix M. Steinkirchner, Franziska Irrgang, Viktoria Kimmerling, Maximilian Käss, Stoyan Popkirov, Elisabeth Schieffer, Michael Gruber, Alexander Dejaco

## Abstract

**Background:** Force production reflects neuromuscular capacity, central regulation, and sensory feedback, making performance-based testing vulnerable when disease-related limitations could be interpreted as reduced effort. Repeated handgrip strength (HGS) testing may quantify neuromuscular fatigability in myalgic encephalomyelitis/chronic fatigue syndrome (ME/CFS), but interpretation of within-subject variability as an effort-validity marker remains controversial. We tested whether the coefficient of variation (CV) distinguishes ME/CFS-related impairment from deliberate submaximal performance and compared it with the sum of residuals (SR), a trajectory-aware metric.

**Methods:** We analyzed three cohorts using the same repeated-HGS protocol: two 10-trial sessions separated by one hour. We included 211 ME/CFS participants and 170 controls, including 28 instructed to perform at 50% perceived maximum force. CV and SR were determined as HGS variability measures and compared by probability-density overlap and receiver operating characteristic analyses. The primary comparison was ME/CFS versus deliberately submaximal controls.

**Results:** CV distributions showed substantial overlap between groups. SR improved group-level separation, increasing non-overlapping fractions from 0.217 to 0.326 in the Jäkel cohort and from 0.180 to 0.361 in the MIRACLE cohort. In ROC analyses, SR consistently yielded higher discrimination than CV across cohorts, sessions, and sex strata. A commonly used 15% CV cutoff classified 42-48% of ME/CFS participants as submaximal, while correctly identifying only 57-64% of deliberately submaximal controls.

**Conclusions:** CV may conflate ME/CFS-related fatigability with irregular force modulation and should not be used as stand-alone evidence of submaximal effort. Trajectory-aware SR partly reduced this bias and indicated that ME/CFS-related force impairment is distinguishable from deliberate submaximal force production at the group level. However, neither metric should be used alone to diagnose ME/CFS or evaluate sincerity of effort.

## Introduction

Voluntary force production reflects the combined influence of neuromuscular capacity, central motor regulation, sensory feedback, pain, and effort allocation, making the interpretation of performance-based testing vulnerable when disease-related limitation is interpreted as reduced effort. This issue is particularly relevant in myalgic encephalomyelitis/chronic fatigue syndrome (ME/CFS), which is characterized by post-exertional malaise (PEM), autonomic dysfunction, pain, cognitive symptoms, and substantial functional impairment [1,2]. Objective markers of neuromuscular fatigability remain limited. Repeated handgrip strength (HGS) testing has been proposed as part of multimodal assessment because it can quantify maximal force, within-session force decline, and recovery across repeated contractions [3,4]. However, analytical approaches remain heterogeneous and interpretation remains debated [5,6,7], limiting broader acceptance, as evidenced by one recent recommendation against using HGS for medicolegal evaluation [8].

The coefficient of variation (CV) of HGS trials has historically been applied as a marker of submaximal effort in occupational and medicolegal contexts [9]. This interpretation rests on the premise that maximal voluntary contractions are relatively consistent, whereas submaximal contractions require graded force control and show greater trial-to-trial variability. More recently, increased CV has been interpreted as compatible with altered higher-order motor control rather than peripheral motor fatigability [5]. Related work has proposed altered effort preference, potentially reflecting protective avoidance behavior and contributing to impaired performance [6]. Although not implying intentional underperformance, these interpretations are sensitive because increased variability has historically supported effort-validity judgments [9].

CV as a stand-alone marker of submaximal effort has been questioned even in its original context [9,10,11]. In ME/CFS, this concern is amplified by the systematic force decline often observed across closely spaced trials, consistent with neuromuscular fatigability and reported to be more pronounced in females and with greater disease severity [3,4,12]. Because CV reflects dispersion around the mean, such decline may increase CV independently of irregular force modulation. CV may therefore conflate systematic fatigability-related decline with variability arising from deliberate force gradation or altered motor control.

We therefore compared CV with the sum of residuals (SR), a trajectory-aware measure derived from individual exponential fits to repeated grip-force series. We analyzed cohorts with ME/CFS and healthy controls (HC) performing maximal contractions, and HC performing deliberately submaximal contractions. We hypothesized that CV would show poor discrimination between ME/CFS-related force impairment and deliberately submaximal force production, whereas SR would improve discrimination by accounting for systematic fatigability-related decline.

## Methods

We analyzed complete datasets from three cohorts using the same HGS protocol: two sessions of ten trials each, one hour apart [3]. Diagnostic criteria, eligibility criteria, and exclusions are detailed in Supplementary Table S1. Cohorts were (i) public Jäkel dataset [3]: Jäkel-ME/CFS (n=105) and Jäkel-HC-max (n=66); (ii) MIRACLE trial clinical cohort: MIR-ME/CFS (n=106) and MIR-HC-max (n=76); (iii) HC-submax: volunteers performing 50% of perceived maximum force (n=28).

HGS was recorded using EH101 dynamometers (Camry Industries Ltd., Hong Kong) in the Jäkel cohort and Baseline® Smedley Digital dynamometers (Fabrication Enterprises Inc., NY, USA) in the MIRACLE and HC-submax cohorts. For each participant and session, CV was calculated from raw grip-force values as 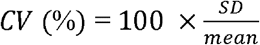. To model systematic force decline, grip-force values were normalized to the individual session-one maximum, and an exponential function was fitted for each session. SR was computed as the sum of distances between observed and fitted force values. For pooled HGS plots, relative HGS values were averaged across participants at each trial.

Discrimination between groups was assessed for CV and SR using probability density functions and receiver operating characteristic (ROC) analysis. The main comparison was pooled ME/CFS versus HC-submax, testing whether disease-related impairment could be distinguished mathematically from deliberate submaximal force gradation. Areas under the curves (AUCs) were compared using DeLong’s test. 95% confidence intervals were estimated by 2000-iteration bootstrapping. Separation of groups was calculated as the non-overlapping fraction (1−OVL). Results were stratified by cohort, session, and sex. Associations with Bell score were assessed by linear regression. Analyses were conducted in R-4.5.1 (R Foundation for Statistical Computing, Vienna, Austria), p<0.05 was considered significant. Medians are presented with interquartile-ranges (IQR).

Ethics approval was obtained (24-3983-101) and all participants gave informed consent. The MIRACLE trial was registered prior to enrollment (DRKS00035953).

## Results

Data from 381 individuals were analyzed: 211 participants with ME/CFS and 170 controls, including 28 HC-submax. Cohort characteristics are provided in Supplementary Tables S1,S2. Mean trajectories showed less within-session decline in HC-max than in ME/CFS or HC-submax, with HC-submax visually strongly overlapping ME/CFS (Figure 1/A1,B1). Sex-stratified curves are shown in Supplementary Figure S1.

**Figure 1.**
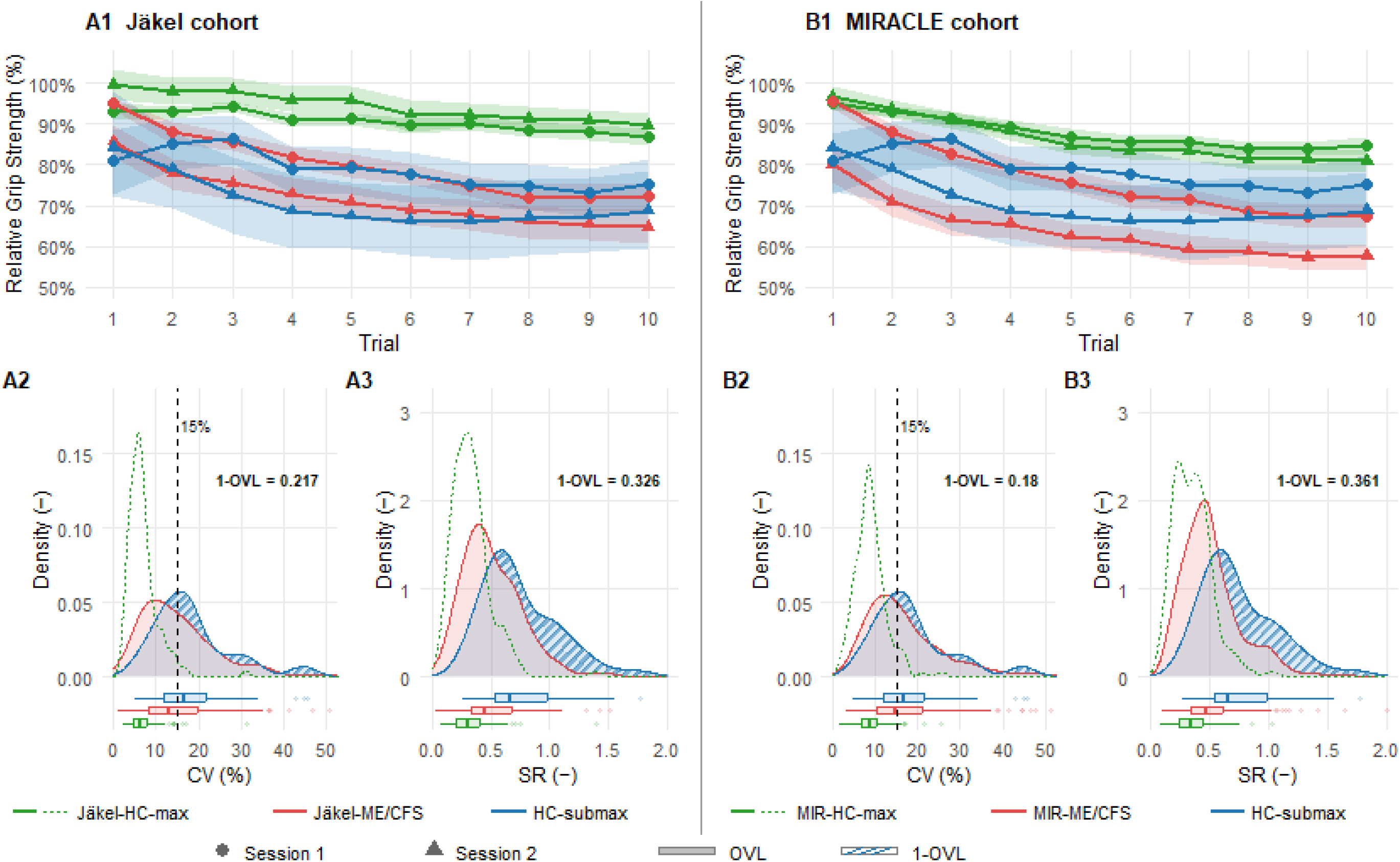
Panels A1 and B1 show mean handgrip strength (HGS) values as percentage of the individual session-one maximum, along two sessions of 10 trials each, one hour apart, for the Jäkel (Jäkel-HC-max, Jäkel-ME/CFS) [3] and MIRACLE (MIR-HC-max, MIR-ME/CFS) cohorts, respectively; each overlaid with a cohort of deliberately submaximal contractions (HC-submax). HGS values are expressed relative to each participant’s individual maximum of session-one. Shaded areas indicate bootstrap 95% confidence intervals of the mean. Panels A2-A3 and B2-B3 show probability density functions of participant-level values of coefficient of variation (CV) and sum of residuals after exponential curve fitting (SR), with data pooled across both HGS sessions. The vertical dashed line in panels A2 and B2 indicates the historically proposed 15% CV threshold for detection of submaximal effort. 1-OVL: non-overlapping area fraction between the ME/CFS and HC-submax distributions; ME/CFS: myalgic encephalomyelitis/chronic fatigue syndrome.

Median CV were 16.69% [11.94-21.68], 14.02% [9.59-20.32], and 7.48% [5.44-9.65] for HC-submax, ME/CFS, and HC-max, respectively; corresponding SR were 0.662 [0.533-0.979], 0.463 [0.335-0.637], and 0.320 [0.222-0.421]. Density plots are shown in Figure 1/A2-B3 and Supplementary Figure S2. ME/CFS vs HC-submax overlap was larger for CV than for SR in both cohorts, with non-overlapping fractions of 0.217 versus 0.326 in Jäkel and 0.180 versus 0.361 in MIRACLE. Across ME/CFS cohorts, lower Bell scores tended to be associated with higher variability (Supplementary Figure S3).

ROC analyses of ME/CFS versus HC-submax (Figure 2; Supplementary Tables S3-S5) showed consistently higher AUCs for SR than CV across cohort, session, and sex. In pooled analyses, highest discrimination occurred in males/session-two (AUC 0.855, 95%-CI 0.738-0.948) and females/session-one (AUC 0.803, 95%-CI 0.671-0.911; Supplementary Figure S4). Applying a previously suggested 15% CV cutoff for submaximal or insincere force application [5,9] assigned 42-48% of ME/CFS participants to the submaximal category, while identifying only 57-64% of participants instructed to perform submaximal contractions, corresponding to balanced accuracy of approximately 55-61% - consistent across cohorts and sessions (Figure 1/A2,B2).

**Figure 2.**
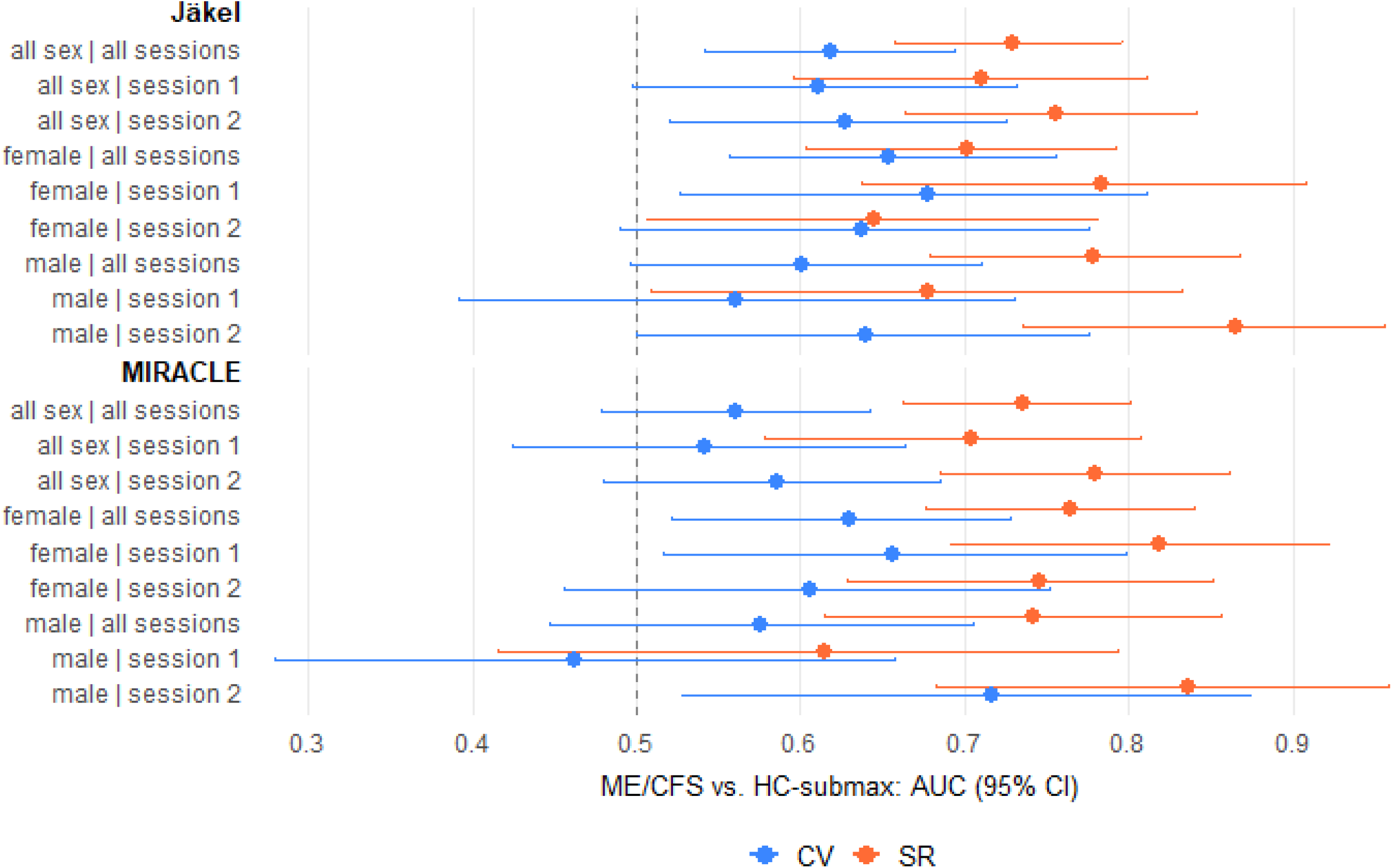
Area under the curve (AUC) estimates from exploratory receiver operating characteristics (ROC) analyses: discrimination capacity of healthy controls executing deliberately submaximal effort (HC-submax) versus participants with myalgic encephalomyelitis/chronic fatigue syndrome (ME/CFS) by intra-individual handgrip-strength variability. Confidence intervals (95% CI) were estimated using 2000 repetition bootstrapping. AUCs are stratified by cohort (Jäkel [3] and MIRACLE), session, and sex. AUC values are shown for the coefficient of variation (CV) method and the sum of residuals (SR) method as variability metrics. The dashed vertical line indicates an AUC of 0.5.

## Discussion

We evaluated CV and SR as metrics to distinguish disease-related fatigability from intentional submaximal force gradation. Consistent with previous studies, both metrics distinguished relatively well HC-submax from HC-max (Figure 1), supporting the concept that deliberate force grading increases variability [9]. CV was more sensitive to distinguish ME/CFS from HC-max, plausibly because CV captures all within-session dispersion, including the systematic force decline characteristic of fatigability in ME/CFS [3,4,12].

The more consequential question was ME/CFS versus HC-submax. Here, CV showed poor discrimination and extensive overlap, highlighting its central limitation: CV cannot distinguish irregular force modulation from systematic force decline. Thus, a smooth but steep force curve can increase CV even without irregular force modulation. In ME/CFS, this may bias interpretation, whereby disease-related fatigability may be misread as other causes of submaximal force gradation or altered higher-order motor control [5,7]. The historically proposed 15% CV cutoff [13] assigned 42-48% of ME/CFS participants to the submaximal category in our dataset, while identifying only 57-64% of HC-submax participants, with balanced accuracy of only 55-61%. Prior literature similarly reported substantial error rates with CV-based effort detection [11,14].

In contrast, SR consistently improved group-level separation between ME/CFS and HC-submax, disentangling fatigability from other factors increasing variability such as intentional force gradation (Figure 1/A3,B3). SR hereby addresses one limitation of CV by fitting individual exponential trajectories and quantifying residual deviations. Exploratory thresholds yielded sensitivity/specificity of 53%/90% in women (AUC=0.803, session-one) and 69%/91% in men (AUC=0.855, session-two) (Supplementary Figure S4). These values remain insufficient for individual effort validation but support further research evaluating SR as a complementary marker. Importantly, our findings demonstrate limitations of CV-based effort-validity testing, but do not exclude central contributions to reduced performance. Altered motor control, altered effort preference, central sensitization, amplified afferent feedback from fatiguing muscle, pain, and protective force regulation to avoid PEM, may all influence motor drive in ME/CFS [5,6,7,15]. These mechanisms could be pathophysiologic or adaptive, not intentional underperformance, and should not be equated with deception or malingering.

Several limitations should be considered. True effort state in ME/CFS could not be directly verified, comparable but different dynamometers were used across cohorts, and no disease-control cohort was available. The HC-submax group was small and performed at 50% perceived maximum force, consistent with previous validity research [9]. However, a comparator mimicking pathological weakness and fatigability may be even better suited in future research.

## Conclusions

In conclusion, HGS may help characterize force, fatigability, and recovery as part of multimodal assessment in ME/CFS. ME/CFS-related variability differed from deliberately submaximal force gradation when systematic decline was modeled, and may in-part reflect peripheral and/or central disease-related mechanisms rather than intentional underperformance. Accordingly, CV should not be used as evidence of submaximal effort. SR reduces this important source of CV-related bias, but neither metric supports definitive individual-level judgments about effort in ME/CFS.

## Data Availability

Data from the MIRACLE cohort (ME/CFS patients, healthy controls performing maximal effort, and healthy controls performing submaximal effort) are provided as supplementary material with this manuscript. Data from the Jaekel cohort were previously published in Jaekel et al. (2021).

## Data Availability Statement

The raw handgrip strength values generated by the authors for the present study are provided as supplementary material. The Jäkel cohort data were obtained from the original publication and remain subject to the data availability conditions of that publication.

## Supplementary Material

Supplement1.pdf contains Supplemental Tables S1-S5, Supplemental Figures S1-S4. Supplement2.csv contains raw handgrip strength values.

## Acknowledgments

We gratefully acknowledge the entire MIRACLE study team at the Universities of Giessen and Marburg and at University Hospital Regensburg for their support and valuable input.

## Disclosure of use of artificial intelligence

AI tools were used for language editing only; authors reviewed all outputs and take full responsibility.

---

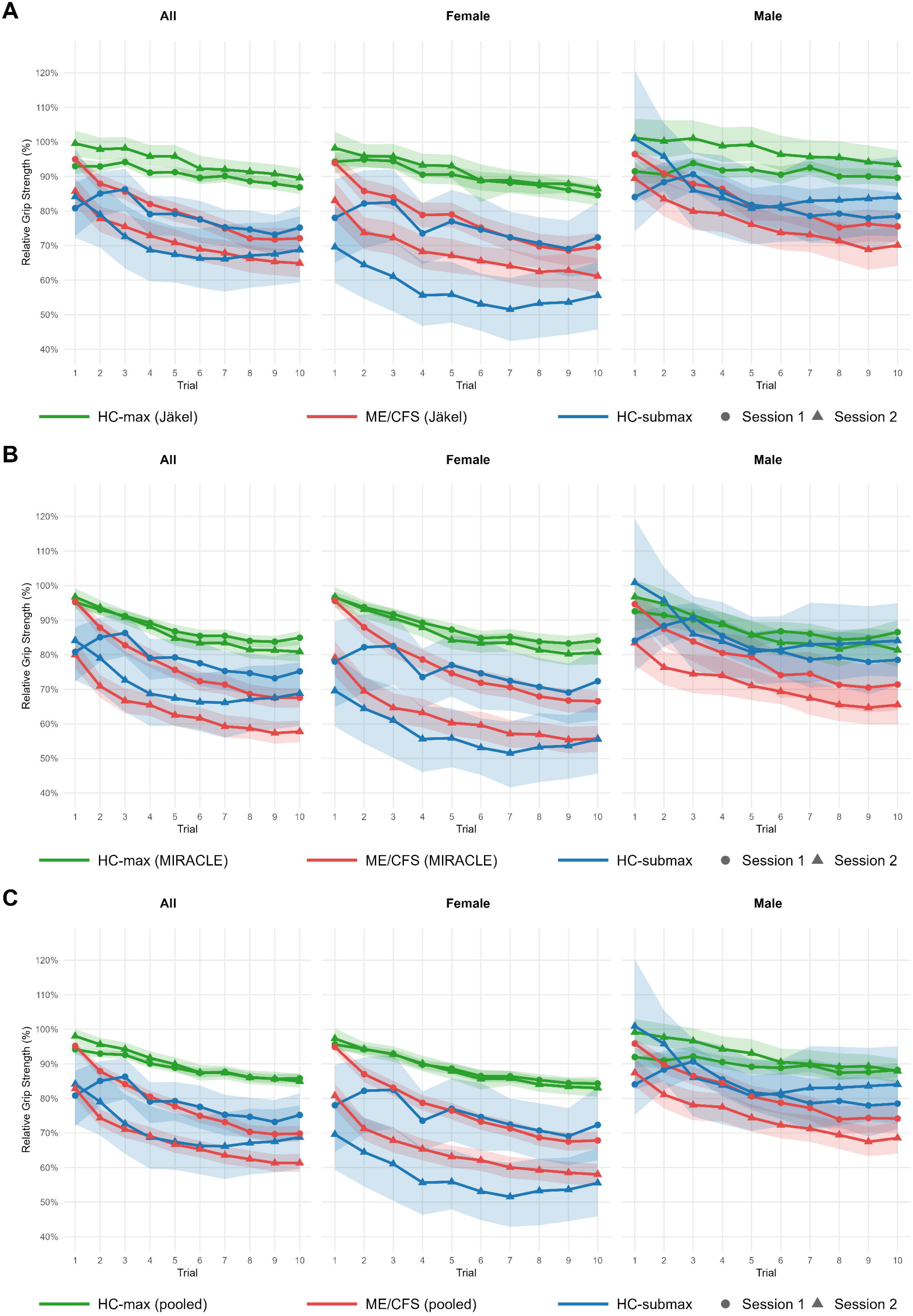

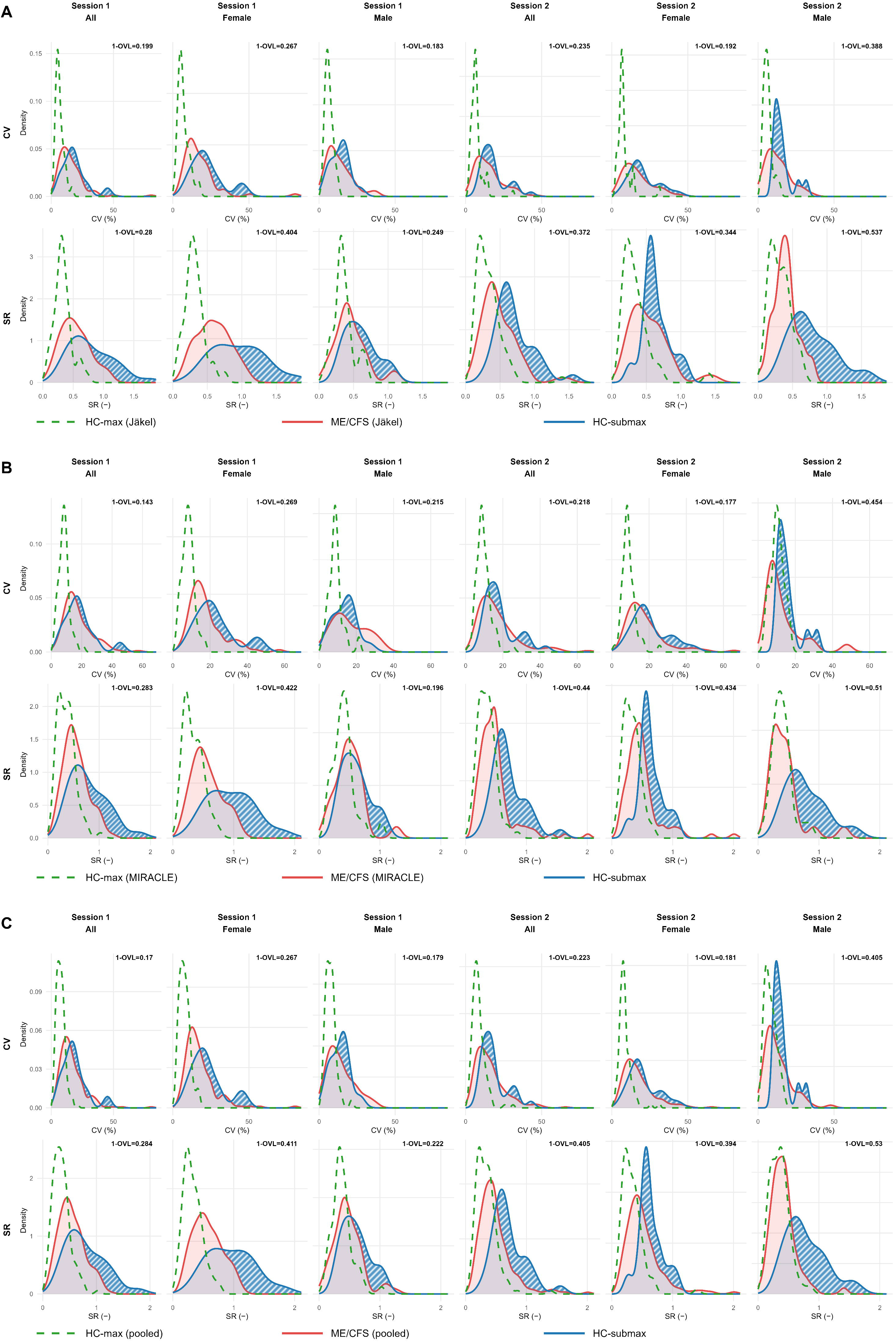

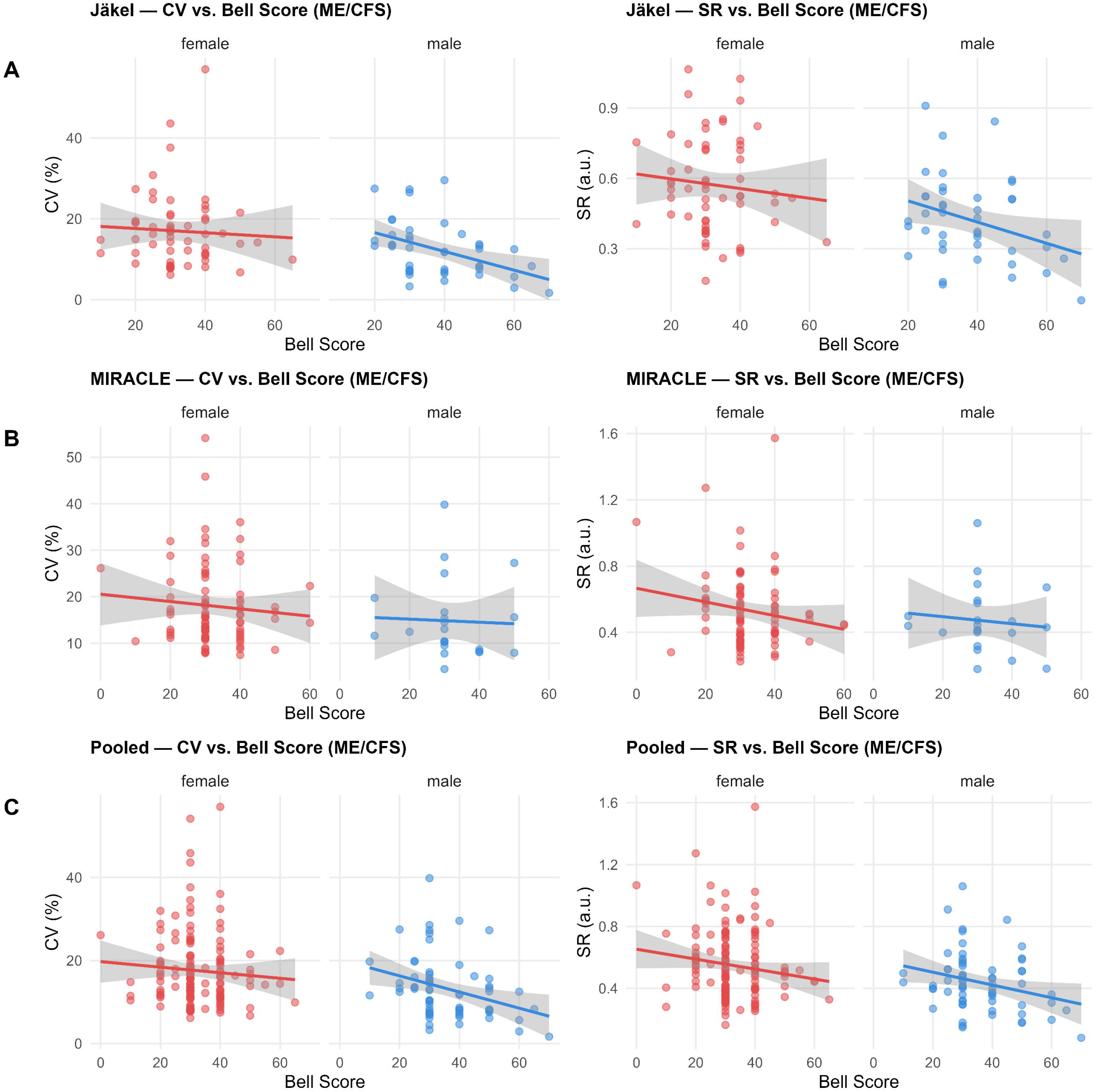

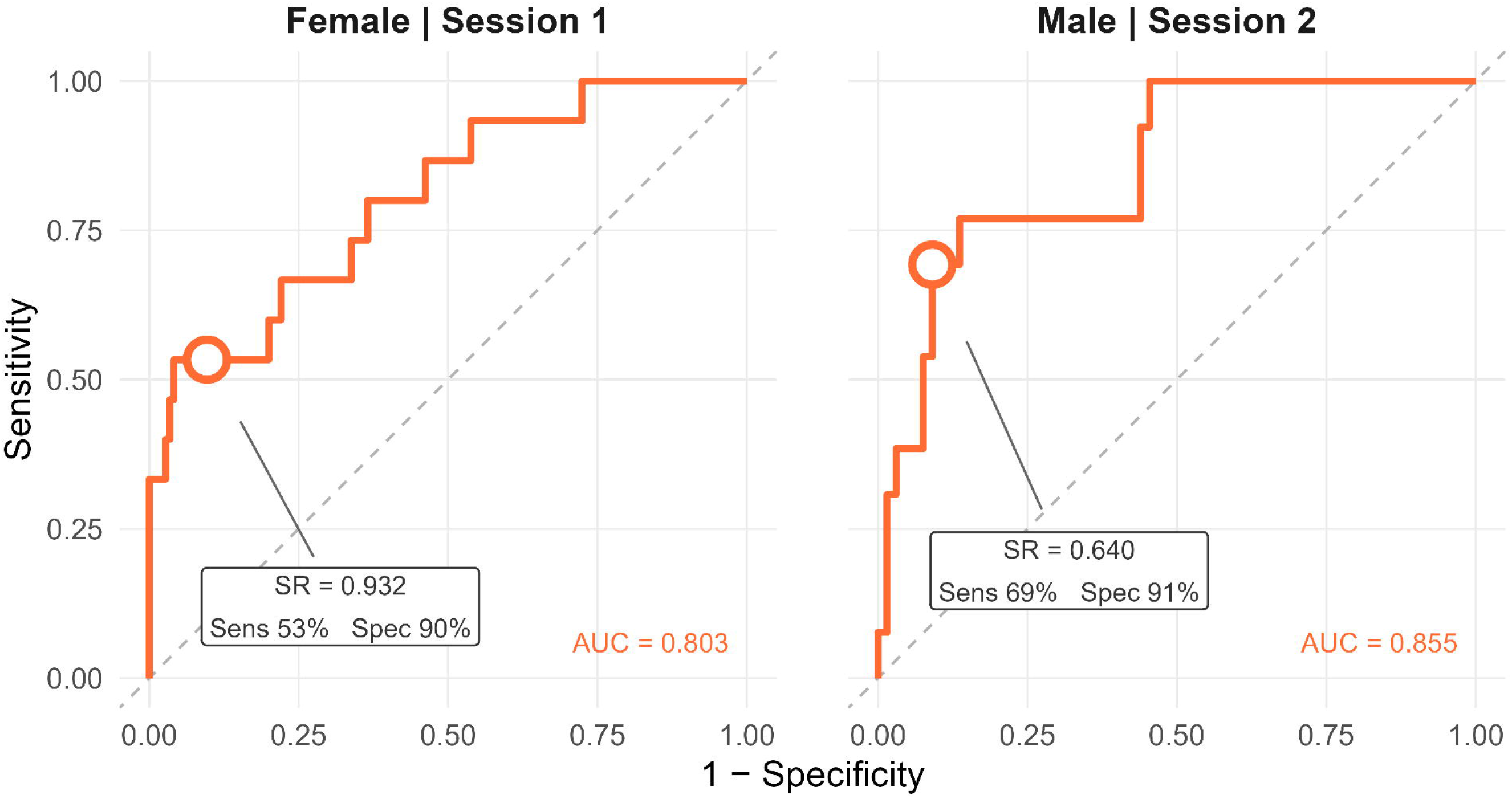

