## Supplement_1 for "Repeated Handgrip Strength Variability in Myalgic Encephalomyelitis/Chronic Fatigue Syndrome: Separating Disease-Related Fatigability from Force Gradation"

**Supplementary Material**

**Supplementary Table S1.** Participant flow, exclusions, and cohort eligibility criteria.

| Cohort / group | n, initial | Diagnostic and eligibility criteria | n, excluded | Reason for exclusion | n, final |
|---|---|---|---|---|---|
| Jäkel–ME/CFS | 105 | Canadian Consensus Criteria; exclusion of other medical or neurological diseases that may cause fatigue [3]. No further exclusion criteria specified. | 0 | — | 105 |
| Jäkel–HC-max | 66 | Self-reported healthy status; age- and sex-matched [3]. | 0 | — | 66 |
| MIR–ME/CFS | 112 | Canadian Consensus Criteria, DSQ-PEM, Disease >6 months; Exclusion: malignancy, surgery within the preceding 6 months, decompensated comorbidities. | 6 | Missing HGS values | 106 |
| MIR–HC-max | 76 | Healthy controls; same exclusion criteria as the MIRACLE ME/CFS group. | 0 | — | 76 |
| HC-submax | 29 | Healthy volunteers without fatigue, neurological, or other HGS-interfering conditions. | 1 | Single missing value | 28 |

HGS, handgrip strength; ME/CFS, myalgic encephalomyelitis/chronic fatigue syndrome; HC-max, healthy controls performing maximal contractions; HC-submax, healthy controls performing deliberately submaximal contractions; MIR, MIRACLE cohort. ME/CFS in all patient groups was diagnosed according to the Canadian Consensus Criteria.

**Supplementary Table S2.** Cohort characteristics.

| Group | n | Female / Male (% F) | Age, median (IQR) | Bell Score, median (IQR) |
|---|---|---|---|---|
| Jäkel cohort – HC-max | 66 | 36 / 30 (54.5%) | 39 (29–50.5) | — |
| Jäkel cohort – ME/CFS | 105 | 61 / 44 (58.1%) | 44 (34–52) | 30 (30–40) |
| MIRACLE cohort – HC-max | 76 | 51 / 25 (67.1%) | 33 (24.8–53) | — |
| MIRACLE cohort – ME/CFS | 106 | 84 / 22 (79.2%) | 45.5 (36–54) | 30 (30–40) |
| Prospective cohort – HC-submax | 28 | 15 / 13 (53.6%) | 29.5 (23.8–36) | — |

Age and Bell Score shown as median (IQR). Bell Score available for ME/CFS patients only (range 0–100; lower scores indicate greater disability). F, female; IQR, interquartile range; ME/CFS, myalgic encephalomyelitis/chronic fatigue syndrome; HC-max, healthy controls performing maximal contractions; HC-submax, healthy controls performing deliberate submaximal contractions.

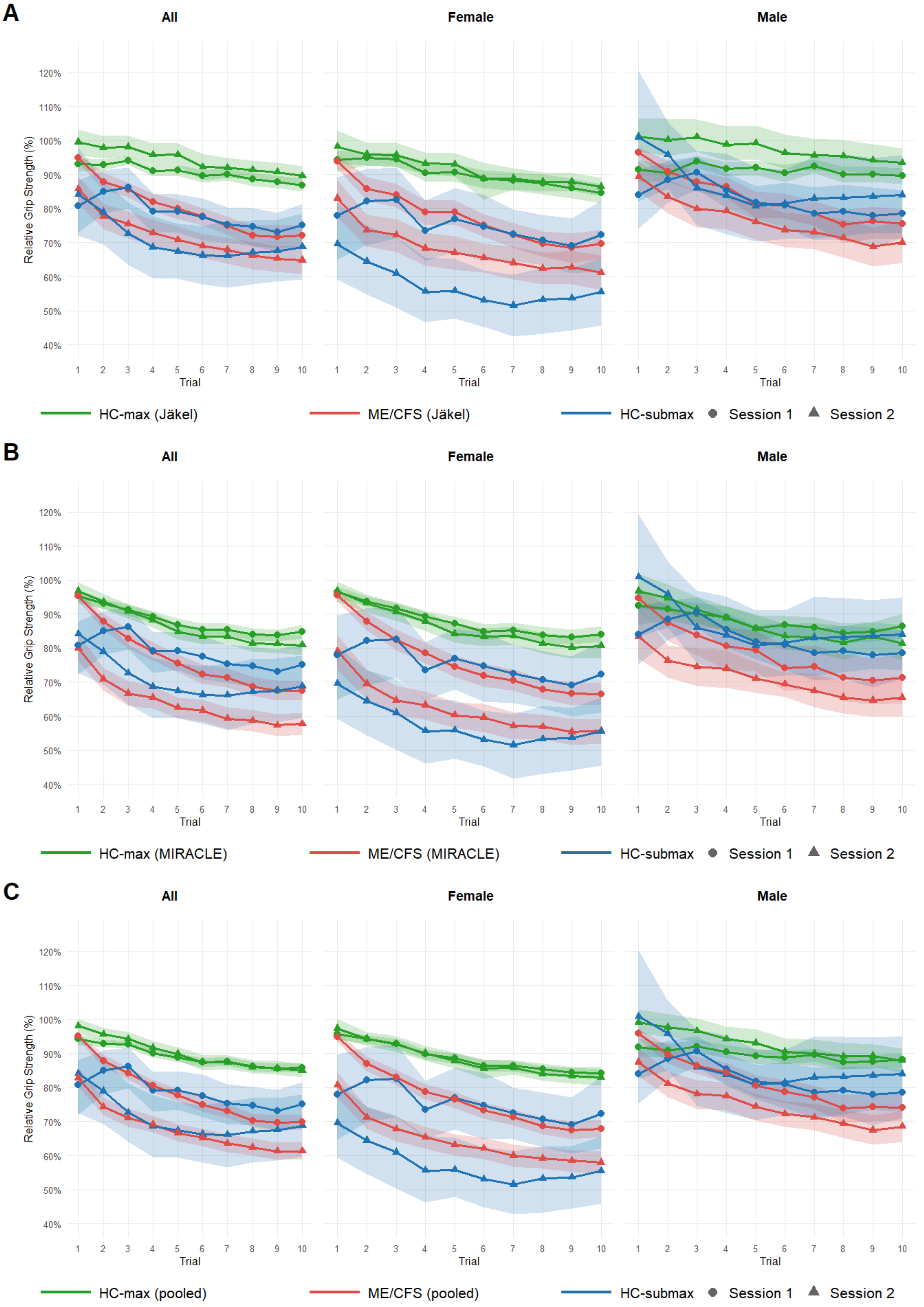


**Supplementary Figure S1.** Grip force curves for Jäkel (A), MIRACLE (B), and pooled (C) cohorts and HC-submax. Mean relative grip strength (± participant-level bootstrapped 95% CI) across 10 trials, stratified by sex. A: Jäkel and HC-submax cohorts. B: MIRACLE and HC-submax cohorts. C: Pooled cohorts (Jäkel and MIRACLE) and HC-submax. CI: confidence interval; HC-max: healthy controls performing maximal contractions; HC-submax: healthy controls performing deliberate submaximal contractions at approximately 50% of perceived maximum force; ME/CFS: participants with myalgic encephalomyelitis/chronic fatigue syndrome asked to perform maximal contractions.

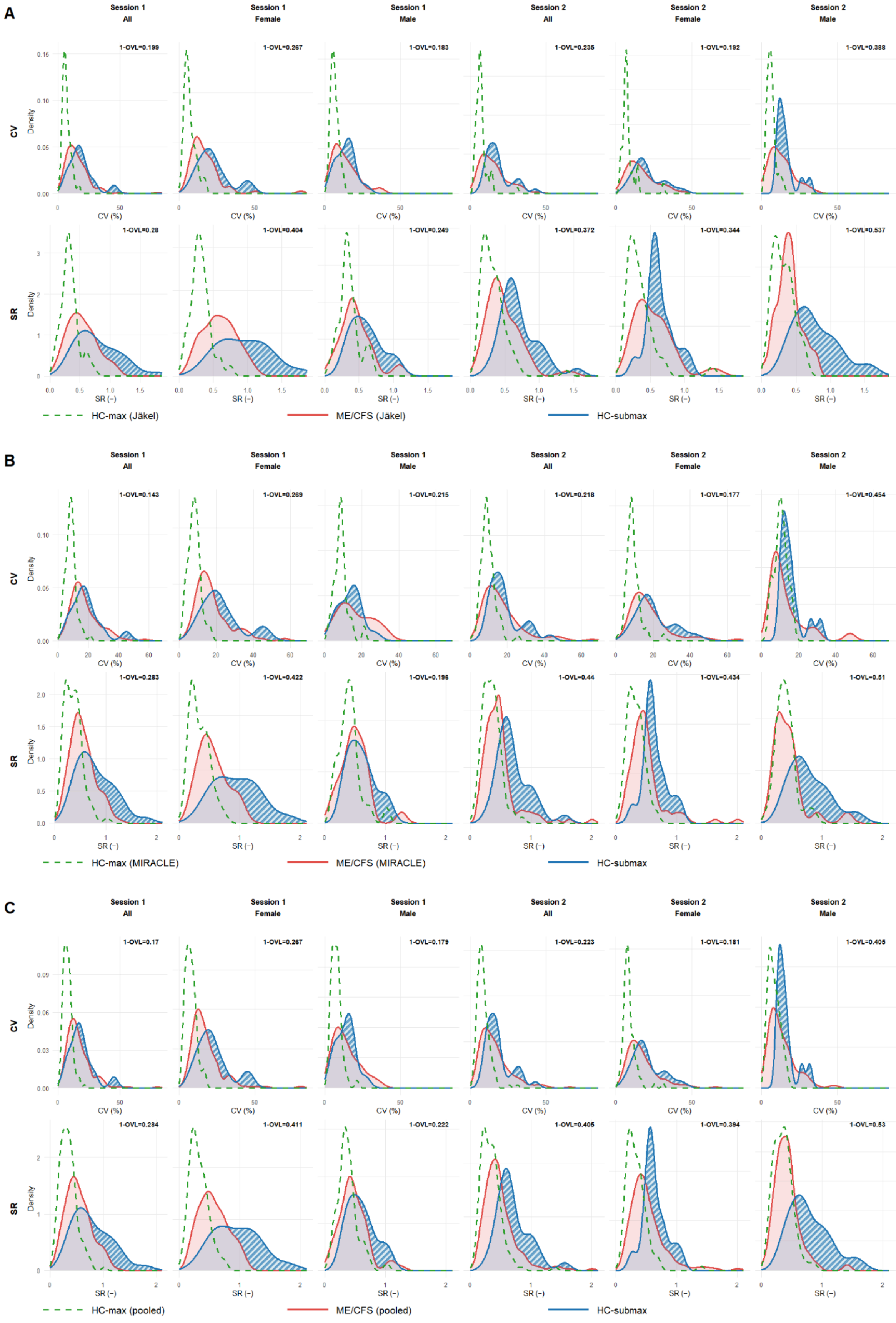


**Supplementary Figure S2.** Probability density functions of CV and SR stratified by session and sex. A: Jäkel and HC-submax cohorts. B: MIRACLE and HC-submax cohorts. C: Pooled cohorts (Jäkel and MIRACLE) and HC-submax. 1-OVL indicates the non-overlapping fraction of the HC-submax and ME/CFS distributions. CV: coefficient of variation (%); HC: healthy control; ME/CFS: myalgic encephalomyelitis/chronic fatigue syndrome; OVL: overlapping coefficient; SR: sum of residuals.

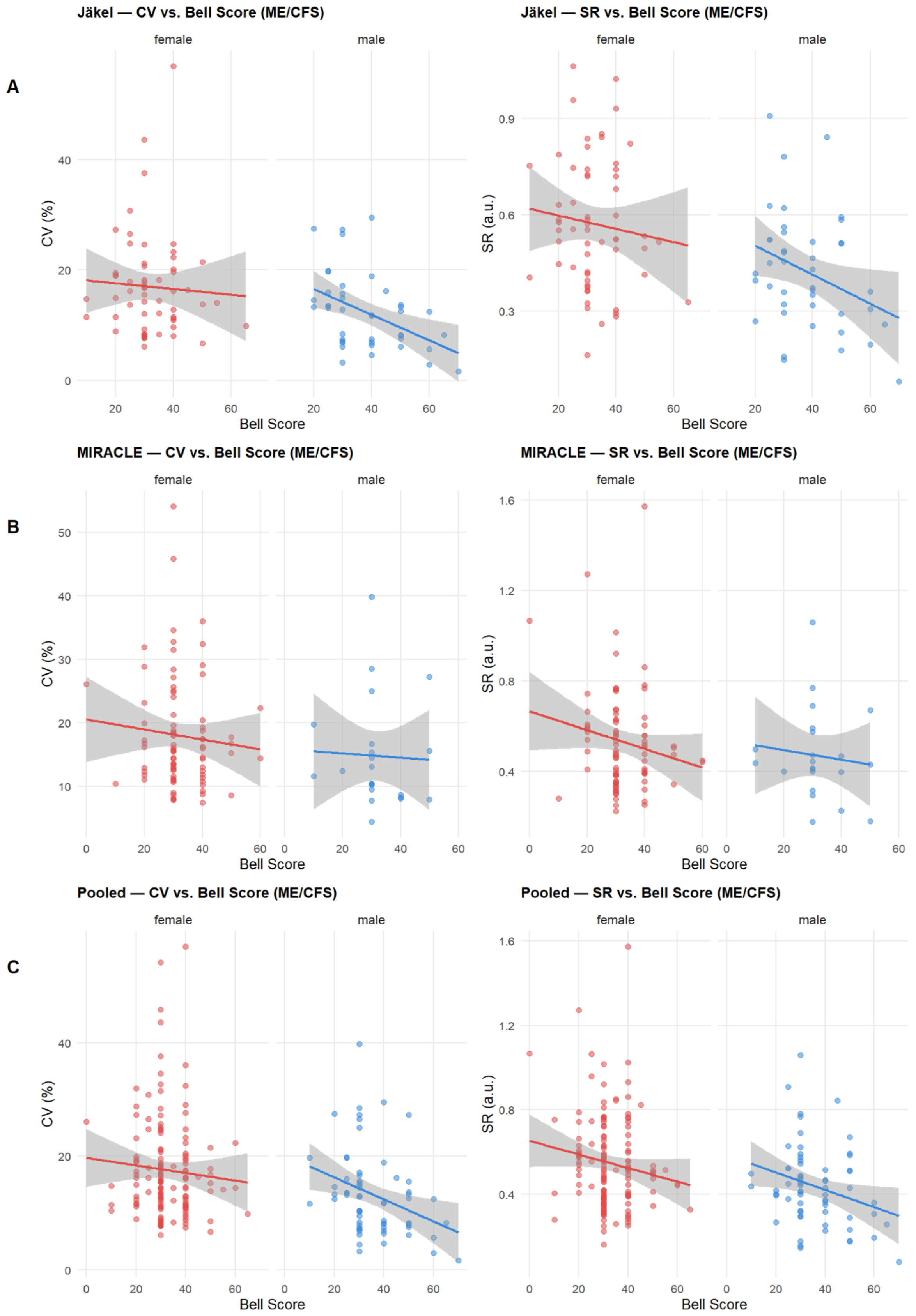


**Supplementary Figure S3.** Association between Bell Score and grip force variability metrics (CV and SR) in ME/CFS patients, stratified by cohort and sex. A: Jäkel cohort. B: MIRACLE cohort. C: Pooled cohorts (Jäkel and MIRACLE). One value per patient (mean over both sessions). Regression lines with 95% confidence bands from linear fits. Lower Bell Score indicates greater disability. CV: coefficient of variation; SR: sum of residuals after exponential trajectory fitting; Bell Score: validated disability scale (0–100).

**Supplementary Table S3:** ROC analysis, Jäkel and HC-submax cohorts.

| Session | Sex | $n_1/n_2$ | Coefficient of Variation (CV, %) | | | Sum of Residuals (SR, -) | | | p DeLong CV vs. SR |
|---|---|---|---|---|---|---|---|---|---|
| | | | G1 median (IQR) | G2 median (IQR) | AUC (95% CI) | G1 median (IQR) | G2 median (IQR) | AUC (95% CI) | |
| **HC-submax (G1) vs. Jäkel-HC (G2)** | | | | | | | | | |
| 1 | all | 28/66 | 17.29 (12.5–21.87) | 5.83 (4.37–8.32) | 0.913 (0.833–0.973) | 0.7 (0.52–1.02) | 0.32 (0.25–0.4) | **0.92 (0.858–0.968)** | 0.7508 |
| 1 | female | 15/36 | 20.46 (15.63–26.32) | 5.87 (4.46–8.58) | 0.965 (0.907–0.998) | 1.01 (0.66–1.24) | 0.29 (0.23–0.37) | **0.97 (0.922–1)** | 0.7826 |
| 1 | male | 13/30 | 15.66 (8.33–17.34) | 5.82 (4.31–7.66) | **0.856 (0.7–0.979)** | 0.56 (0.45–0.73) | 0.32 (0.29–0.42) | 0.849 (0.715–0.949) | 0.8703 |
| 2 | all | 28/66 | 15.97 (11.9–19.51) | 6.39 (5.01–7.76) | **0.946 (0.899–0.982)** | 0.63 (0.55–0.84) | 0.3 (0.2–0.4) | 0.913 (0.85–0.964) | 0.0896 |
| 2 | female | 15/36 | 17.84 (15.11–25.55) | 6.63 (5.57–7.87) | **0.941 (0.863–0.996)** | 0.58 (0.55–0.73) | 0.3 (0.21–0.4) | 0.889 (0.781–0.974) | 0.0418* |
| 2 | male | 13/30 | 13.75 (11.78–16.08) | 5.94 (4.16–7.26) | **0.962 (0.897–1)** | 0.68 (0.61–0.98) | 0.3 (0.19–0.4) | 0.951 (0.877–0.995) | 0.7218 |
| pooled | all | 56/132 | 16.69 (11.94–21.68) | 6.11 (4.53–7.87) | **0.929 (0.882–0.967)** | 0.66 (0.53–0.98) | 0.31 (0.21–0.40) | 0.913 (0.867–0.950) | 0.3123 |
| pooled | female | 30/72 | 18.25 (14.73–27.57) | 6.39 (4.78–8.10) | **0.950 (0.906–0.985)** | 0.70 (0.57–1.02) | 0.30 (0.21–0.39) | 0.929 (0.869–0.974) | 0.1707 |
| pooled | male | 26/60 | 14.27 (11.16–17.05) | 5.86 (4.19–7.66) | **0.916 (0.829–0.979)** | 0.65 (0.45–0.88) | 0.32 (0.21–0.40) | 0.898 (0.827–0.958) | 0.4775 |
| **HC-submax (G1) vs. Jäkel-ME/CFS (G2)** | | | | | | | | | |
| 1 | all | 28/105 | 17.29 (12.5–21.87) | 12.94 (8.76–19.9) | 0.611 (0.501–0.725) | 0.7 (0.52–1.02) | 0.51 (0.35–0.7) | **0.71 (0.599–0.813)** | 0.0170* |
| 1 | female | 15/61 | 20.46 (15.63–26.32) | 14.73 (10.95–20.39) | 0.678 (0.523–0.817) | 1.01 (0.66–1.24) | 0.57 (0.41–0.74) | **0.783 (0.642–0.907)** | 0.0853 |
| 1 | male | 13/44 | 15.66 (8.33–17.34) | 11.43 (6.97–17.11) | 0.561 (0.381–0.731) | 0.56 (0.45–0.73) | 0.41 (0.31–0.57) | **0.678 (0.516–0.829)** | 0.0467* |
| 2 | all | 28/105 | 15.97 (11.9–19.51) | 12.8 (8.16–18.43) | 0.627 (0.521–0.724) | 0.63 (0.55–0.84) | 0.44 (0.32–0.63) | **0.755 (0.663–0.841)** | 0.0094** |
| 2 | female | 15/61 | 17.84 (15.11–25.55) | 14.88 (9.91–21.85) | 0.637 (0.49–0.773) | 0.58 (0.55–0.73) | 0.47 (0.34–0.68) | **0.645 (0.498–0.78)** | 0.9136 |
| 2 | male | 13/44 | 13.75 (11.78–16.08) | 11.29 (6.69–17.74) | 0.64 (0.488–0.771) | 0.68 (0.61–0.98) | 0.38 (0.3–0.46) | **0.865 (0.747–0.96)** | 0.0010** |
| pooled | all | 56/210 | 16.69 (11.94–21.68) | 12.91 (8.41–19.60) | 0.618 (0.541–0.697) | 0.66 (0.53–0.98) | 0.45 (0.33–0.67) | **0.729 (0.656–0.795)** | 0.0005*** |
| pooled | female | 30/122 | 18.25 (14.73–27.57) | 14.80 (10.15–21.18) | 0.654 (0.549–0.753) | 0.70 (0.57–1.02) | 0.53 (0.36–0.72) | **0.701 (0.599–0.796)** | 0.3212 |
| pooled | male | 26/88 | 14.27 (11.16–17.05) | 11.29 (6.81–17.63) | 0.601 (0.490–0.708) | 0.65 (0.45–0.88) | 0.40 (0.31–0.53) | **0.778 (0.676–0.864)** | 0.0000*** |
| **Jäkel-ME/CFS (G1) vs. Jäkel-HC (G2)** | | | | | | | | | |
| 1 | all | 105/66 | 12.94 (8.76–19.9) | 5.83 (4.37–8.32) | **0.847 (0.784–0.901)** | 0.51 (0.35–0.7) | 0.32 (0.25–0.4) | 0.758 (0.68–0.831) | 0.0012** |
| 1 | female | 61/36 | 14.73 (10.95–20.39) | 5.87 (4.46–8.58) | **0.89 (0.816–0.948)** | 0.57 (0.41–0.74) | 0.29 (0.23–0.37) | 0.829 (0.746–0.905) | 0.0745 |
| 1 | male | 44/30 | 11.43 (6.97–17.11) | 5.82 (4.31–7.66) | **0.773 (0.664–0.879)** | 0.41 (0.31–0.57) | 0.32 (0.29–0.42) | 0.65 (0.52–0.776) | 0.0066** |
| 2 | all | 105/66 | 12.8 (8.16–18.43) | 6.39 (5.01–7.76) | **0.815 (0.747–0.875)** | 0.44 (0.32–0.63) | 0.3 (0.2–0.4) | 0.705 (0.625–0.781) | 0.0002*** |
| 2 | female | 61/36 | 14.88 (9.91–21.85) | 6.63 (5.57–7.87) | **0.83 (0.737–0.909)** | 0.47 (0.34–0.68) | 0.3 (0.21–0.4) | 0.742 (0.636–0.841) | 0.0160* |
| 2 | male | 44/30 | 11.29 (6.69–17.74) | 5.94 (4.16–7.26) | **0.798 (0.69–0.888)** | 0.38 (0.3–0.46) | 0.3 (0.19–0.4) | 0.643 (0.511–0.768) | 0.0008*** |

| | | | | | | | | | |
|---|---|---|---|---|---|---|---|---|---|
| pooled | all | 210/132 | 12.91 (8.41–19.60) | 6.11 (4.53–7.87) | **0.832 (0.790–0.873)** | 0.45 (0.33–0.67) | 0.31 (0.21–0.40) | 0.732 (0.676–0.782) | 0.0000*** |
| pooled | female | 122/72 | 14.80 (10.15–21.18) | 6.39 (4.78–8.10) | **0.863 (0.808–0.912)** | 0.53 (0.36–0.72) | 0.30 (0.21–0.39) | 0.788 (0.722–0.849) | 0.0018** |
| pooled | male | 88/60 | 11.29 (6.81–17.63) | 5.86 (4.19–7.66) | **0.789 (0.708–0.855)** | 0.40 (0.31–0.53) | 0.32 (0.21–0.40) | 0.649 (0.557–0.737) | 0.0000*** |

G1 and G2 as indicated in each block header. Median (IQR) of raw CV and SR per group. AUC from ROC analysis with bootstrapped 95% CI. p (DeLong): DeLong test comparing AUC of CV versus SR. Bold AUC indicates the higher-discriminating method. AUC, area under the curve; CI, confidence interval; CV, coefficient of variation (%); HC, healthy control; IQR, interquartile range; ME/CFS, myalgic encephalomyelitis/chronic fatigue syndrome; ROC, receiver operating characteristic; SR, sum of residuals (a.u.). *p<0.05, **p<0.01, ***p<0.001.

**Supplementary Table S4:** ROC analysis, MIRACLE and HC-submax cohorts.

| Session | Sex | $n_1$/$n_2$ | Coefficient of Variation (CV, %) | | | Sum of Residuals (SR, -) | | | p DeLong CV vs. SR |
|---|---|---|---|---|---|---|---|---|---|
| | | | G1 median (IQR) | G2 median (IQR) | AUC (95% CI) | G1 median (IQR) | G2 median (IQR) | AUC (95% CI) | |
| **HC-submax (G1) vs. MIRACLE-HC (G2)** | | | | | | | | | |
| 1 | all | 28/76 | 17.29 (12.5–21.87) | 8.14 (6.06–9.97) | 0.861 (0.76–0.947) | 0.7 (0.52–1.02) | 0.35 (0.22–0.45) | **0.883 (0.804–0.948)** | 0.5166 |
| 1 | female | 15/51 | 20.46 (15.63–26.32) | 8.06 (6.06–9.94) | 0.95 (0.873–0.997) | 1.01 (0.66–1.24) | 0.28 (0.21–0.44) | **0.957 (0.894–0.996)** | 0.8559 |
| 1 | male | 13/25 | 15.66 (8.33–17.34) | 8.61 (7.17–9.97) | 0.742 (0.545–0.92) | 0.56 (0.45–0.73) | 0.4 (0.32–0.47) | **0.757 (0.572–0.902)** | 0.8035 |
| 2 | all | 28/76 | 15.97 (11.9–19.51) | 8.83 (7.37–11.23) | 0.878 (0.8–0.945) | 0.63 (0.55–0.84) | 0.33 (0.24–0.42) | **0.916 (0.846–0.967)** | 0.1454 |
| 2 | female | 15/51 | 17.84 (15.11–25.55) | 8.59 (7.12–10.65) | 0.906 (0.791–0.988) | 0.58 (0.55–0.73) | 0.31 (0.22–0.4) | **0.925 (0.831–0.99)** | 0.4055 |
| 2 | male | 13/25 | 13.75 (11.78–16.08) | 10.16 (8.64–12.36) | 0.797 (0.637–0.92) | 0.68 (0.61–0.98) | 0.37 (0.27–0.46) | **0.883 (0.757–0.975)** | 0.2059 |
| pooled | all | 56/152 | 16.69 (11.94–21.68) | 8.62 (6.79–10.55) | 0.871 (0.810–0.930) | 0.66 (0.53–0.98) | 0.33 (0.23–0.45) | **0.898 (0.849–0.940)** | 0.2212 |
| pooled | female | 30/102 | 18.25 (14.73–27.57) | 8.28 (6.75–10.16) | 0.928 (0.863–0.978) | 0.70 (0.57–1.02) | 0.31 (0.21–0.41) | **0.937 (0.880–0.978)** | 0.7113 |
| pooled | male | 26/50 | 14.27 (11.16–17.05) | 9.24 (7.54–11.41) | 0.781 (0.655–0.891) | 0.65 (0.45–0.88) | 0.38 (0.29–0.47) | **0.822 (0.717–0.911)** | 0.3189 |
| **HC-submax (G1) vs. MIRACLE-ME/CFS (G2)** | | | | | | | | | |
| 1 | all | 28/106 | 17.29 (12.5–21.87) | 14.85 (11–22.45) | 0.542 (0.424–0.659) | 0.7 (0.52–1.02) | 0.5 (0.38–0.69) | **0.704 (0.591–0.812)** | 0.0001*** |
| 1 | female | 15/84 | 20.46 (15.63–26.32) | 15.06 (11.61–20.46) | 0.656 (0.51–0.792) | 1.01 (0.66–1.24) | 0.52 (0.38–0.72) | **0.818 (0.695–0.928)** | 0.0030** |
| 1 | male | 13/22 | 15.66 (8.33–17.34) | 14.62 (10.17–24.23) | 0.462 (0.266–0.654) | 0.56 (0.45–0.73) | 0.49 (0.38–0.62) | **0.615 (0.402–0.797)** | 0.0309* |
| 2 | all | 28/106 | 15.97 (11.9–19.51) | 14.23 (9.49–21.02) | 0.585 (0.478–0.689) | 0.63 (0.55–0.84) | 0.43 (0.32–0.53) | **0.78 (0.687–0.863)** | 0.0000*** |
| 2 | female | 15/84 | 17.84 (15.11–25.55) | 15.33 (11.04–21.45) | 0.606 (0.46–0.747) | 0.58 (0.55–0.73) | 0.45 (0.33–0.55) | **0.746 (0.626–0.859)** | 0.0177* |
| 2 | male | 13/22 | 13.75 (11.78–16.08) | 9.51 (6.4–14.74) | 0.717 (0.531–0.874) | 0.68 (0.61–0.98) | 0.39 (0.27–0.51) | **0.836 (0.682–0.958)** | 0.0836 |
| pooled | all | 56/212 | 16.69 (11.94–21.68) | 14.72 (10.39–21.22) | 0.561 (0.474–0.641) | 0.66 (0.53–0.98) | 0.47 (0.34–0.62) | **0.736 (0.668–0.801)** | 0.0000*** |
| pooled | female | 30/168 | 18.25 (14.73–27.57) | 15.18 (11.13–21.22) | 0.630 (0.522–0.734) | 0.70 (0.57–1.02) | 0.48 (0.35–0.63) | **0.764 (0.682–0.845)** | 0.0014** |
| pooled | male | 26/44 | 14.27 (11.16–17.05) | 10.94 (7.29–21.25) | 0.575 (0.440–0.705) | 0.65 (0.45–0.88) | 0.44 (0.28–0.54) | **0.742 (0.614–0.857)** | 0.0004*** |
| **MIRACLE-ME/CFS (G1) vs. MIRACLE-HC (G2)** | | | | | | | | | |
| 1 | all | 106/76 | 14.85 (11–22.45) | 8.14 (6.06–9.97) | **0.862 (0.805–0.911)** | 0.5 (0.38–0.69) | 0.35 (0.22–0.45) | 0.739 (0.661–0.804) | 0.0002*** |
| 1 | female | 84/51 | 15.06 (11.61–20.46) | 8.06 (6.06–9.94) | **0.891 (0.833–0.938)** | 0.52 (0.38–0.72) | 0.28 (0.21–0.44) | 0.777 (0.699–0.853) | 0.0029** |
| 1 | male | 22/25 | 14.62 (10.17–24.23) | 8.61 (7.17–9.97) | **0.769 (0.611–0.907)** | 0.49 (0.38–0.62) | 0.4 (0.32–0.47) | 0.645 (0.476–0.798) | 0.0718 |
| 2 | all | 106/76 | 14.23 (9.49–21.02) | 8.83 (7.37–11.23) | **0.757 (0.684–0.824)** | 0.43 (0.32–0.53) | 0.33 (0.24–0.42) | 0.67 (0.592–0.746) | 0.0265* |
| 2 | female | 84/51 | 15.33 (11.04–21.45) | 8.59 (7.12–10.65) | **0.831 (0.758–0.895)** | 0.45 (0.33–0.55) | 0.31 (0.22–0.4) | 0.722 (0.633–0.808) | 0.0188* |
| 2 | male | 22/25 | 9.51 (6.4–14.74) | 10.16 (8.64–12.36) | 0.489 (0.316–0.662) | 0.39 (0.27–0.51) | 0.37 (0.27–0.46) | **0.522 (0.353–0.685)** | 0.8441 |

| | | | | | | | | | |
|---|---|---|---|---|---|---|---|---|---|
| pooled | all | 212/152 | 14.72 (10.39–21.22) | 8.62 (6.79–10.55) | **0.811 (0.767–0.853)** | 0.47 (0.34–0.62) | 0.33 (0.23–0.45) | 0.705 (0.651–0.757) | 0.0000*** |
| pooled | female | 168/102 | 15.18 (11.13–21.22) | 8.28 (6.75–10.16) | **0.861 (0.816–0.906)** | 0.48 (0.35–0.63) | 0.31 (0.21–0.41) | 0.750 (0.689–0.806) | 0.0002*** |
| pooled | male | 44/50 | 10.94 (7.29–21.25) | 9.24 (7.54–11.41) | **0.643 (0.529–0.757)** | 0.44 (0.28–0.54) | 0.38 (0.29–0.47) | 0.584 (0.465–0.699) | 0.2064 |

G1 and G2 as indicated in each block header. Median (IQR) of raw CV and SR per group. AUC from ROC analysis with bootstrapped 95% CI. p (DeLong): DeLong test comparing AUC of CV versus SR. Bold AUC indicates the higher-discriminating method. AUC, area under the curve; CI, confidence interval; CV, coefficient of variation (%); HC, healthy control; IQR, interquartile range; ME/CFS, myalgic encephalomyelitis/chronic fatigue syndrome; ROC, receiver operating characteristic; SR, sum of residuals (a.u.). *p<0.05, **p<0.01, ***p<0.001.

**Supplementary Table S5:** ROC analysis, pooled (Jäkel and MIRACLE) and HC-submax cohorts.

| Session | Sex | $n_1/n_2$ | Coefficient of Variation (CV, %) | | | Sum of Residuals (SR, -) | | | p DeLong CV vs. SR |
|---|---|---|---|---|---|---|---|---|---|
| | | | G1 median (IQR) | G2 median (IQR) | AUC (95% CI) | G1 median (IQR) | G2 median (IQR) | AUC (95% CI) | |
| **HC-submax (G1) vs. HC-max (G2)** | | | | | | | | | |
| 1 | all | 28/142 | 17.29 (12.5–21.87) | 7.15 (5–9.44) | 0.885 (0.792–0.955) | 0.7 (0.52–1.02) | 0.32 (0.23–0.43) | **0.9 (0.837–0.953)** | 0.5655 |
| 1 | female | 15/87 | 20.46 (15.63–26.32) | 7.13 (4.9–9.77) | 0.956 (0.894–0.995) | 1.01 (0.66–1.24) | 0.29 (0.21–0.41) | **0.962 (0.91–0.995)** | 0.8292 |
| 1 | male | 13/55 | 15.66 (8.33–17.34) | 7.17 (5.35–9.2) | 0.804 (0.634–0.952) | 0.56 (0.45–0.73) | 0.35 (0.29–0.45) | **0.807 (0.677–0.916)** | 0.9504 |
| 2 | all | 28/142 | 15.97 (11.9–19.51) | 7.61 (5.84–10.2) | 0.91 (0.85–0.958) | 0.63 (0.55–0.84) | 0.31 (0.22–0.41) | **0.914 (0.854–0.959)** | 0.7970 |
| 2 | female | 15/87 | 17.84 (15.11–25.55) | 7.62 (6.14–9.36) | **0.92 (0.831–0.985)** | 0.58 (0.55–0.73) | 0.31 (0.21–0.4) | 0.91 (0.818–0.975) | 0.5570 |
| 2 | male | 13/55 | 13.75 (11.78–16.08) | 7.47 (5.13–10.79) | 0.887 (0.792–0.957) | 0.68 (0.61–0.98) | 0.35 (0.23–0.43) | **0.92 (0.839–0.982)** | 0.3564 |
| pooled | all | 56/284 | 16.69 (11.94–21.68) | 7.48 (5.44–9.65) | 0.898 (0.846–0.941) | 0.66 (0.53–0.98) | 0.32 (0.22–0.42) | **0.905 (0.862–0.941)** | 0.6658 |
| pooled | female | 30/174 | 18.25 (14.73–27.57) | 7.54 (5.55–9.71) | **0.938 (0.887–0.979)** | 0.70 (0.57–1.02) | 0.30 (0.21–0.41) | 0.934 (0.883–0.972) | 0.8166 |
| pooled | male | 26/110 | 14.27 (11.16–17.05) | 7.32 (5.14–9.54) | 0.855 (0.762–0.936) | 0.65 (0.45–0.88) | 0.35 (0.25–0.45) | **0.864 (0.786–0.931)** | 0.7346 |
| **HC-submax (G1) vs. ME/CFS (G2)** | | | | | | | | | |
| 1 | all | 28/211 | 17.29 (12.5–21.87) | 14.48 (10.09–20.37) | 0.576 (0.462–0.691) | 0.7 (0.52–1.02) | 0.51 (0.37–0.7) | **0.707 (0.596–0.808)** | 0.0005*** |
| 1 | female | 15/145 | 20.46 (15.63–26.32) | 14.94 (11.16–20.39) | 0.665 (0.514–0.797) | 1.01 (0.66–1.24) | 0.54 (0.39–0.73) | **0.803 (0.671–0.911)** | 0.0092** |
| 1 | male | 13/66 | 15.66 (8.33–17.34) | 12.3 (7.46–19.38) | 0.528 (0.382–0.679) | 0.56 (0.45–0.73) | 0.42 (0.32–0.59) | **0.657 (0.5–0.797)** | 0.0154* |
| 2 | all | 28/211 | 15.97 (11.9–19.51) | 13.35 (8.96–19.83) | 0.606 (0.51–0.694) | 0.63 (0.55–0.84) | 0.44 (0.32–0.58) | **0.768 (0.683–0.845)** | 0.0001*** |
| 2 | female | 15/145 | 17.84 (15.11–25.55) | 15.13 (10.25–21.49) | 0.619 (0.473–0.743) | 0.58 (0.55–0.73) | 0.45 (0.33–0.62) | **0.703 (0.588–0.798)** | 0.1407 |
| 2 | male | 13/66 | 13.75 (11.78–16.08) | 10.43 (6.57–17.28) | 0.666 (0.543–0.784) | 0.68 (0.61–0.98) | 0.38 (0.27–0.48) | **0.855 (0.738–0.948)** | 0.0010** |
| pooled | all | 56/422 | 16.69 (11.94–21.68) | 14.02 (9.59–20.32) | 0.589 (0.514–0.659) | 0.66 (0.53–0.98) | 0.46 (0.34–0.64) | **0.732 (0.668–0.796)** | 0.0000*** |
| pooled | female | 30/290 | 18.25 (14.73–27.57) | 15.03 (10.83–21.21) | 0.640 (0.541–0.737) | 0.70 (0.57–1.02) | 0.49 (0.35–0.69) | **0.737 (0.650–0.817)** | 0.0164* |
| pooled | male | 26/132 | 14.27 (11.16–17.05) | 11.01 (6.93–17.87) | 0.592 (0.485–0.688) | 0.65 (0.45–0.88) | 0.41 (0.30–0.53) | **0.766 (0.669–0.855)** | 0.0000*** |
| **ME/CFS (G1) vs. HC-max (G2)** | | | | | | | | | |
| 1 | all | 211/142 | 14.48 (10.09–20.37) | 7.15 (5–9.44) | **0.845 (0.802–0.883)** | 0.51 (0.37–0.7) | 0.32 (0.23–0.43) | 0.75 (0.699–0.8) | 0.0000*** |
| 1 | female | 145/87 | 14.94 (11.16–20.39) | 7.13 (4.9–9.77) | **0.887 (0.84–0.925)** | 0.54 (0.39–0.73) | 0.29 (0.21–0.41) | 0.805 (0.747–0.859) | 0.0011** |
| 1 | male | 66/55 | 12.3 (7.46–19.38) | 7.17 (5.35–9.2) | **0.744 (0.654–0.823)** | 0.42 (0.32–0.59) | 0.35 (0.29–0.45) | 0.636 (0.537–0.732) | 0.0054** |
| 2 | all | 211/142 | 13.35 (8.96–19.83) | 7.61 (5.84–10.2) | **0.78 (0.732–0.826)** | 0.44 (0.32–0.58) | 0.31 (0.22–0.41) | 0.687 (0.629–0.741) | 0.0002*** |
| 2 | female | 145/87 | 15.13 (10.25–21.49) | 7.62 (6.14–9.36) | **0.828 (0.775–0.88)** | 0.45 (0.33–0.62) | 0.31 (0.21–0.4) | 0.732 (0.666–0.797) | 0.0015** |
| 2 | male | 66/55 | 10.43 (6.57–17.28) | 7.47 (5.13–10.79) | **0.686 (0.596–0.779)** | 0.38 (0.27–0.48) | 0.35 (0.23–0.43) | 0.59 (0.487–0.689) | 0.0171* |
| pooled | all | 422/284 | 14.02 (9.59–20.32) | 7.48 (5.44–9.65) | **0.814 (0.781–0.843)** | 0.46 (0.34–0.64) | 0.32 (0.22–0.42) | 0.718 (0.680–0.754) | 0.0000*** |
| pooled | female | 290/174 | 15.03 (10.83–21.21) | 7.54 (5.55–9.71) | **0.859 (0.823–0.892)** | 0.49 (0.35–0.69) | 0.30 (0.21–0.41) | 0.768 (0.725–0.810) | 0.0000*** |

| | | | | | | | | | |
|---|---|---|---|---|---|---|---|---|---|
| pooled | male | 132/110 | 11.01 (6.93–17.87) | 7.32 (5.14–9.54) | **0.717 (0.651–0.779)** | 0.41 (0.30–0.53) | 0.35 (0.25–0.45) | 0.611 (0.543–0.681) | 0.0002*** |

G1 and G2 as indicated in each block header. Median (IQR) of raw CV and SR per group. AUC from ROC analysis with bootstrapped 95% CI. p (DeLong): DeLong test comparing AUC of CV versus SR. Bold AUC indicates the higher-discriminating method. AUC, area under the curve; CI, confidence interval; CV, coefficient of variation (%); HC, healthy control; IQR, interquartile range; ME/CFS, myalgic encephalomyelitis/chronic fatigue syndrome; ROC, receiver operating characteristic; SR, sum of residuals (a.u.). *p<0.05, **p<0.01, ***p<0.001.

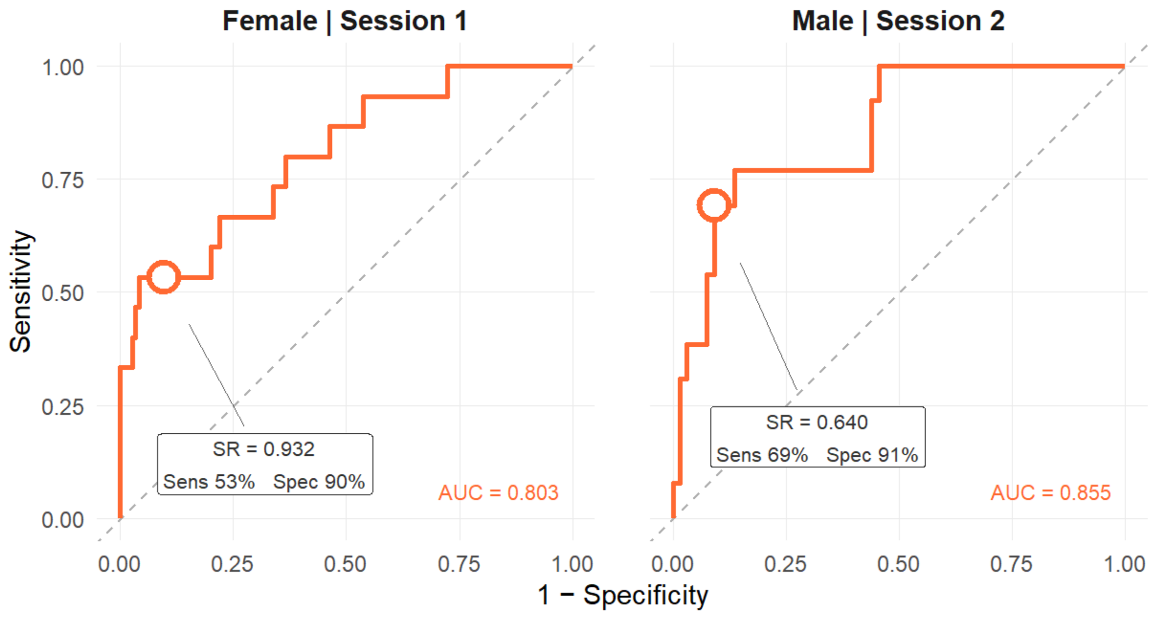


**Supplementary Figure S4:** Receiver operating characteristic (ROC) curve for the sum of residuals (SR) distinguishing deliberately submaximal force production (HC-submax) from ME/CFS-related grip force impairment, stratified by sex. Data pooled across both cohorts (Jäkel and MIRACLE) and both sessions. Circle: SR threshold at ≥90% specificity, selected to minimise false-positive misclassification of ME/CFS participants as submaximal - the primary risk in clinical and medicolegal contexts. At this threshold, ≥90% of ME/CFS participants are correctly classified as non-submaximal. AUC estimated by 2000-repetition bootstrapping. SR: sum of absolute residuals after individual exponential trajectory fitting to session-1-normalised grip force values; AUC: area under the ROC curve.
