## Supplement_2 for "Repeated Handgrip Strength Variability in Myalgic Encephalomyelitis/Chronic Fatigue Syndrome: Separating Disease-Related Fatigability from Force Gradation"

| Group | sex | Age_Class | HGS_1_1 | HGS_2_1 | HGS_3_1 | HGS_4_1 | HGS_5_1 | HGS_6_1 | HGS_7_1 | HGS_8_1 | HGS_9_1 | HGS_10_1 | HGS_1_2 | HGS_2_2 | HGS_3_2 | HGS_4_2 | HGS_5_2 | HGS_6_2 | HGS_7_2 | HGS_8_2 | HGS_9_2 | HGS_10_2 |
|---|---|---|---|---|---|---|---|---|---|---|---|---|---|---|---|---|---|---|---|---|---|---|
| MIRACLE-MECFS | m | 40-49 | 56.2 | 49.8 | 50.7 | 48.8 | 49.4 | 49.1 | 45.4 | 45.3 | 45.1 | 44.7 | 56.6 | 53 | 51.7 | 49.5 | 50.2 | 47.5 | 46.4 | 46.8 | 43.2 | 43.8 |
| MIRACLE-MECFS | m | 30-39 | 54.7 | 49.7 | 31.5 | 32.9 | 32 | 31.9 | 26.9 | 23 | 24.9 | 26 | 46.9 | 44.5 | 37.7 | 36.3 | 38.8 | 38.6 | 27.2 | 28 | 31.8 | 30.7 |
| MIRACLE-MECFS | m | 30-39 | 48.5 | 44.5 | 45.2 | 39.9 | 36.1 | 37.9 | 34.6 | 32.4 | 33.5 | 33.4 | 37.7 | 34.1 | 38.9 | 36.3 | 38.1 | 36.6 | 32.8 | 33.6 | 35.4 | 33.4 |
| MIRACLE-MECFS | m | 50-59 | 44.9 | 39.8 | 33.4 | 32.5 | 28.5 | 23.5 | 28.3 | 30.4 | 26.3 | 22.3 | 25.7 | 25.7 | 27.3 | 30 | 28.9 | 28.4 | 28.4 | 29.8 | 25.6 | 29.6 |
| MIRACLE-MECFS | m | 50-59 | 44.2 | 36.4 | 38.1 | 34.7 | 34.7 | 31.2 | 25.3 | 25 | 23.1 | 26.2 | 43.8 | 41.4 | 38.5 | 41.3 | 40.4 | 34.2 | 40.2 | 39.3 | 32.7 | 34.11 |
| MIRACLE-MECFS | m | 30-39 | 44.2 | 29.8 | 37.9 | 33.2 | 32.4 | 28.7 | 27.1 | 31 | 26.7 | 29.2 | 38.3 | 37.1 | 39.4 | 38.5 | 36 | 37.1 | 35.7 | 34.3 | 31.6 | 35 |
| MIRACLE-MECFS | m | 30-39 | 42.2 | 44.8 | 46.7 | 44.7 | 42.1 | 37.9 | 38.5 | 39.6 | 33.7 | 33.9 | 44.6 | 35.7 | 41.3 | 37.2 | 31.7 | 32.7 | 29.4 | 32.9 | 29.1 | 30.6 |
| MIRACLE-MECFS | m | 50-59 | 40.9 | 41.2 | 41.4 | 43.3 | 40 | 40.9 | 39.3 | 33.2 | 34.3 | 37.3 | 28.7 | 33 | 32.6 | 32.9 | 29.6 | 27 | 28.6 | 27.7 | 30.3 | 30.7 |
| MIRACLE-MECFS | f | 40-49 | 40.6 | 37.2 | 37.6 | 35.6 | 34.6 | 37.5 | 31.3 | 30.1 | 29.1 | 28.2 | 32.1 | 35.5 | 28.7 | 26.7 | 25.6 | 27.5 | 23.4 | 23 | 23.4 | 27.1 |
| MIRACLE-MECFS | m | 40-49 | 40.6 | 40.2 | 39.9 | 39.4 | 39.3 | 37.2 | 37.65 | 38.8 | 37.6 | 37.2 | 36.1 | 36.3 | 36.2 | 33.4 | 28.5 | 29 | 28.1 | 26.1 | 28.5 | 27.4 |
| MIRACLE-MECFS | m | 50-59 | 40.4 | 41.7 | 41.8 | 41.5 | 41.2 | 41.3 | 41.1 | 39.5 | 38.9 | 37.4 | 27.8 | 26.8 | 26.3 | 25.6 | 30.1 | 26.5 | 26.3 | 25.3 | 25.7 | 25.9 |
| MIRACLE-MECFS | m | 30-39 | 39.8 | 45.1 | 28.7 | 42.5 | 32.8 | 29.7 | 32.1 | 24.7 | 24.4 | 22 | 43.5 | 39.5 | 31.4 | 41.7 | 24.7 | 24.6 | 23.5 | 24.2 | 21.6 | 20.8 |
| MIRACLE-MECFS | m | 40-49 | 36.7 | 37.3 | 25.9 | 25.1 | 21.5 | 20.1 | 18 | 17.8 | 18.6 | 20 | 31.4 | 24.1 | 22.4 | 18.8 | 17.8 | 17.1 | 15.7 | 14.9 | 16.9 | 15.2 |
| MIRACLE-MECFS | m | 50-59 | 36.5 | 35.3 | 33.2 | 29.5 | 32.6 | 30 | 28.3 | 24.1 | 23.8 | 27.4 | 30.1 | 30.5 | 32.2 | 30.2 | 28 | 26.9 | 26.9 | 27.5 | 27.7 | 27.8 |
| MIRACLE-MECFS | f | 40-49 | 36.5 | 34.5 | 33.2 | 29.9 | 30.3 | 30.1 | 27.1 | 27.5 | 24.3 | 26.8 | 29.8 | 30.6 | 28.5 | 27.1 | 26.9 | 26.9 | 23.4 | 24 | 21.2 | 21.4 |
| MIRACLE-MECFS | m | 50-59 | 36 | 43.7 | 43 | 41 | 42.1 | 35 | 36.4 | 33.3 | 35.2 | 33.4 | 32.5 | 34.2 | 31.8 | 31.2 | 33.8 | 31.7 | 30.5 | 30.7 | 26.8 | 30.9 |
| MIRACLE-MECFS | f | 50-59 | 35 | 34.2 | 28.8 | 26.4 | 22.4 | 25.5 | 24 | 23.4 | 22 | 22.1 | 21.5 | 18.1 | 18.7 | 21.1 | 17.5 | 20.2 | 17.1 | 22.8 | 17.1 | 18.6 |
| MIRACLE-MECFS | m | 20-29 | 34.6 | 25.2 | 27.8 | 24.9 | 27 | 25 | 26.7 | 24.9 | 24.9 | 22.8 | 26.3 | 26.2 | 18.6 | 12.5 | 19.6 | 23.5 | 25 | 17.8 | 20.2 | 18.5 |
| MIRACLE-MECFS | f | 50-59 | 34 | 32.5 | 31.1 | 29.1 | 28.4 | 29.2 | 27.5 | 26.6 | 27.2 | 24.1 | 31.8 | 29.4 | 28.4 | 26.5 | 29.4 | 25.2 | 27 | 22.2 | 22.1 | 21.2 |
| MIRACLE-MECFS | m | 60-69 | 32.3 | 24.7 | 21.5 | 22 | 40.9 | 29 | 27.6 | 28.7 | 41.2 | 42.5 | 31.2 | 27.5 | 32.1 | 32.2 | 32.1 | 31.2 | 31.7 | 29.4 | 30.4 | 29.4 |
| MIRACLE-MECFS | f | 20-29 | 31.1 | 26.4 | 26 | 25.5 | 26.3 | 24.8 | 24.9 | 21.9 | 23.7 | 23.5 | 29.5 | 21.9 | 22.4 | 20.1 | 18.3 | 19.4 | 18.2 | 17.8 | 16.8 | 16.8 |
| MIRACLE-MECFS | f | 40-49 | 30.9 | 26.7 | 26.3 | 28.9 | 30.1 | 29.6 | 28.6 | 28 | 25.1 | 26.6 | 32 | 30.6 | 30.3 | 29.1 | 27.6 | 29.2 | 26.3 | 24 | 24.1 | 23.9 |
| MIRACLE-MECFS | f | 20-29 | 30.6 | 28.3 | 25.9 | 23.7 | 24.3 | 22.6 | 21.7 | 22.2 | 22.1 | 21.1 | 24.1 | 27.5 | 18.7 | 20.4 | 22.6 | 18.1 | 20 | 16.9 | 19.3 | 18 |
| MIRACLE-MECFS | f | 50-59 | 30.2 | 28.5 | 27.4 | 24.7 | 25.3 | 25.1 | 24.9 | 25.6 | 24.7 | 24.2 | 30.7 | 28.2 | 25.7 | 24.2 | 25.9 | 22.9 | 23.6 | 24.6 | 23 | 22.4 |
| MIRACLE-MECFS | f | 40-49 | 29.8 | 28.5 | 24.6 | 24.7 | 19.7 | 17.9 | 20.4 | 17.1 | 17.3 | 15.5 | 23.1 | 20.7 | 18.8 | 17.2 | 15.3 | 14.9 | 17 | 15.2 | 17 | 12.8 |
| MIRACLE-MECFS | f | 40-49 | 29.3 | 25.3 | 23.2 | 21.9 | 18.7 | 17.3 | 19.6 | 20.5 | 17.8 | 16.1 | 21.4 | 23.6 | 17.1 | 19.4 | 18.8 | 15 | 17 | 20.4 | 15.3 | 15.8 |
| MIRACLE-MECFS | m | 50-59 | 29 | 20 | 22 | 20.3 | 21.7 | 19.7 | 18.6 | 19.2 | 19.8 | 18.3 | 24.7 | 22.2 | 19.2 | 20.8 | 19.7 | 20.2 | 18.3 | 18.6 | 18.5 | 18.8 |
| MIRACLE-MECFS | f | 40-49 | 28.5 | 26 | 20.4 | 20.3 | 20.7 | 20 | 21.8 | 21 | 19.8 | 19.7 | 19.4 | 15.7 | 12.9 | 14.2 | 13.3 | 13.2 | 10.9 | 10.5 | 8.5 | 11.6 |
| MIRACLE-MECFS | f | 20-29 | 28.4 | 28.1 | 23.9 | 22 | 19.8 | 22.2 | 24.1 | 22.9 | 18 | 20.6 | 20.6 | 21.8 | 20.2 | 22.7 | 18.3 | 18.2 | 19.6 | 15.4 | 15.5 | 18.4 |
| MIRACLE-MECFS | f | 50-59 | 28.1 | 27.3 | 29.3 | 27.7 | 27.2 | 27.5 | 26.2 | 25.9 | 25.8 | 25.5 | 29.9 | 29.2 | 28.3 | 26.7 | 25.2 | 25.4 | 23.3 | 22.8 | 22.5 | 22.4 |
| MIRACLE-MECFS | f | 50-59 | 27.7 | 25 | 23.7 | 17.5 | 25.3 | 26 | 24.2 | 25.6 | 24.6 | 25.3 | 22.5 | 17.8 | 17 | 20.1 | 18.7 | 21.6 | 18.8 | 20.3 | 20.8 | 22.4 |
| MIRACLE-MECFS | f | 30-39 | 27.5 | 26.9 | 28.3 | 25.1 | 24.9 | 20.3 | 22.2 | 25.9 | 25 | 20.4 | 22.9 | 18.6 | 15.7 | 17.3 | 13 | 13.5 | 17.3 | 18.9 | 21.3 | 17.8 |
| MIRACLE-MECFS | f | 50-59 | 27.5 | 25.5 | 25.5 | 24.5 | 24.1 | 22.5 | 22.9 | 21 | 25 | 23.2 | 23 | 26.2 | 26.3 | 24.7 | 24.2 | 25 | 26.6 | 23.8 | 20.7 | 23.9 |
| MIRACLE-MECFS | f | 40-49 | 27.4 | 25.1 | 21.8 | 22.2 | 26.1 | 22 | 25.2 | 20.5 | 21.6 | 17.3 | 20.9 | 21.9 | 17.4 | 19.4 | 18.7 | 20.6 | 20.1 | 16.8 | 20.2 | 25.2 |
| MIRACLE-MECFS | f | 30-39 | 27.2 | 30.1 | 28.3 | 29.1 | 23 | 22.8 | 20.4 | 20.1 | 19.2 | 19.2 | 28 | 24.7 | 24.1 | 22.4 | 24.6 | 22.8 | 22.6 | 22.7 | 21.5 | 20.5 |
| MIRACLE-MECFS | f | 20-29 | 27.2 | 21.4 | 17.9 | 17 | 16.6 | 18.2 | 22.3 | 18.9 | 20.7 | 19.3 | 25.3 | 22.3 | 21.4 | 18.5 | 17.3 | 16.4 | 17.9 | 13.1 | 12.4 | 15.1 |
| MIRACLE-MECFS | f | 20-29 | 26.8 | 20.3 | 24.2 | 22.9 | 21.4 | 20.9 | 18.2 | 16.6 | 19.1 | 19 | 21.5 | 14.5 | 16.2 | 14.6 | 12.5 | 11.5 | 11.3 | 11.8 | 11 | 10.7 |
| MIRACLE-MECFS | f | 40-49 | 26.6 | 24.5 | 18.3 | 18.7 | 16.8 | 17.5 | 13.1 | 13.7 | 13.4 | 15.2 | 19.8 | 15.7 | 15.5 | 14.8 | 14.7 | 13.9 | 13.5 | 15.4 | 16.7 | 17.6 |
| MIRACLE-MECFS | f | 20-29 | 26.5 | 16.8 | 17.5 | 14.5 | 13.1 | 11.1 | 11.6 | 13.6 | 9.6 | 15.5 | 21.7 | 22.4 | 18.2 | 14.2 | 12.8 | 13.6 | 13.3 | 13.7 | 13.7 | 13.2 |
| MIRACLE-MECFS | f | 50-59 | 26.4 | 23.6 | 21.7 | 19.3 | 19.1 | 19.5 | 18.9 | 15.6 | 16.3 | 17.7 | 25.3 | 23 | 17.4 | 18.5 | 16.5 | 16.1 | 14.9 | 16.4 | 9.3 | 11.6 |
| MIRACLE-MECFS | m | 40-49 | 26.4 | 27.1 | 24.4 | 20.2 | 19.8 | 14 | 16.7 | 16.7 | 14.2 | 15.3 | 15.7 | 17.6 | 15.3 | 18.9 | 16.4 | 17.6 | 16.3 | 14.7 | 12.6 | 11.9 |
| MIRACLE-MECFS | m | 50-59 | 26.1 | 22.2 | 23.8 | 22.8 | 18 | 19 | 22.3 | 23.2 | 20.4 | 21.4 | 22.5 | 23.3 | 21.3 | 20.6 | 19.5 | 18.7 | 20.4 | 17.9 | 19 | 20.3 |
| MIRACLE-MECFS | f | 20-29 | 25.5 | 23.2 | 21.8 | 20.4 | 18.8 | 17.9 | 19.4 | 18.1 | 16.2 | 18.4 | 22.9 | 19.6 | 19.7 | 16.3 | 14.5 | 15.1 | 14.8 | 14.3 | 14.5 | 14 |
| MIRACLE-MECFS | f | 20-29 | 25 | 22.9 | 19.3 | 20.6 | 18.4 | 16.6 | 17.3 | 16.1 | 15.7 | 17.2 | 21.1 | 16.6 | 14 | 15.6 | 15.8 | 11.6 | 13.4 | 17 | 12.3 | 17.9 |
| MIRACLE-MECFS | f | 30-39 | 24.7 | 11.1 | 7.3 | 10.2 | 8.1 | 7 | 8 | 7.4 | 5.7 | 7.1 | 15 | 5.2 | 6.8 | 3.8 | 5.4 | 4.7 | 4.1 | 5.7 | 5.6 | 9.5 |
| MIRACLE-MECFS | m | 40-49 | 24.4 | 24.3 | 27.9 | 28.1 | 28 | 29.6 | 29.1 | 26.3 | 26.4 | 27.9 | 24.4 | 24.2 | 22.8 | 23.1 | 25.7 | 22.5 | 19.7 | 19.8 | 20 | 24.2 |
| MIRACLE-MECFS | f | 50-59 | 24.2 | 19.8 | 21.3 | 17.7 | 20.9 | 16.4 | 16.5 | 17.3 | 19.1 | 16.9 | 22.2 | 21.1 | 21.4 | 16.9 | 15.6 | 19 | 19 | 16.7 | 17.4 | 16.3 |
| MIRACLE-MECFS | f | 40-49 | 24.1 | 25.2 | 24.2 | 20.4 | 19.8 | 18.9 | 20 | 19.6 | 20.2 | 14.8 | 22.4 | 22.9 | 18.5 | 16.3 | 19.9 | 16.2 | 17.3 | 15.1 | 15.8 | 15.1 |
| MIRACLE-MECFS | f | 40-49 | 24 | 14.4 | 12.2 | 11.5 | 10.2 | 9.1 | 10.7 | 8.3 | 10.7 | 8.5 | 10 | 8.2 | 8 | 9.1 | 9.1 | 9.4 | 7.6 | 6.7 | 8.9 | 9.6 |
| MIRACLE-MECFS | f | 30-39 | 23.8 | 26.5 | 27.1 | 24.4 | 26.5 | 24.5 | 22.5 | 21.3 | 22 | 20.9 | 23.5 | 24.8 | 22.8 | 22.9 | 21.7 | 20.9 | 22.2 | 19.7 | 22.2 | 20.8 |
| MIRACLE-MECFS | f | 50-59 | 23.6 | 25.9 | 18.6 | 16.7 | 17.5 | 16.8 | 16 | 15.6 | 15.1 | 15.6 | 20 | 18.9 | 15.1 | 14.8 | 13.9 | 14.6 | 15.3 | 16.8 | 14.5 | 14 |
| MIRACLE-MECFS | f | 30-39 | 23.5 | 19.3 | 18 | 16.8 | 17 | 16.3 | 18.1 | 16.1 | 16.1 | 16.2 | 17.6 | 20.4 | 18.7 | 18.5 | 17.8 | 16.6 | 16.6 | 16.4 | 17.5 | 14.3 |
| MIRACLE-MECFS | f | 50-59 | 23.3 | 21.2 | 22.9 | 21.6 | 19.9 | 18.7 | 15.6 | 19.2 | 17.9 | 17.9 | 21.5 | 21.9 | 22.6 | 21.7 | 18.6 | 21 | 18 | 18.9 | 18.1 | 18.4 |
| MIRACLE-MECFS | f | 30-39 | 23 | 23.9 | 19.6 | 17.8 | 19.3 | 19.1 | 17.1 | 18.9 | 15.8 | 16.1 | 25.4 | 21.6 | 19.7 | 17.6 | 17.1 | 18.5 | 16 | 15.2 | 16.2 | 18.1 |
| MIRACLE-MECFS | f | 20-29 | 22.7 | 19.3 | 19.3 | 16.9 | 17.7 | 16 | 13.7 | 14.1 | 15.9 | 14.8 | 16 | 11.9 | 12.8 | 13.6 | 10.3 | 11.2 | 7.9 | 9.3 | 11.2 | 7.4 |
| MIRACLE-MECFS | f | 30-39 | 22.7 | 22.4 | 22.4 | 20.1 | 19.6 | 19.8 | 20 | 18.7 | 18 | 18.7 | 21.1 | 23.1 | 23.4 | 22.8 | 23.1 | 23.8 | 22.2 | 20.3 | 19.7 | 19.6 |
| MIRACLE-MECFS | f | 50-59 | 22.4 | 19.3 | 19.8 | 19.3 | 18.6 | 19.1 | 16.4 | 16.2 | 19.4 | 13.2 | 16.2 | 17.8 | 12.7 | 18 | 12.9 | 13.8 | 14.9 | 13.5 | 15.4 | 15.6 |
| MIRACLE-MECFS | f | 50-59 | 22.1 | 20.7 | 18.9 | 19.3 | 19.2 | 19 | 18.8 | 17.8 | 16.4 | 15.6 | 17.5 | 17.1 | 17.1 | 16.2 | 14 | 14.6 | 15.2 | 15.8 | 15.6 | 16.8 |
| MIRACLE-MECFS | f | 40-49 | 22.1 | 15.9 | 11.8 | 16.2 | 12 | 13.5 | 14.1 | 13.4 | 11.7 | 11.5 | 7.2 | 9.6 | 7.2 | 8.8 | 10.1 | 8 | 9 | 10.4 | 6.9 | 8.1 |

| | | | | | | | | | | | | | | | | | | | | | | |
|---|---|---|---|---|---|---|---|---|---|---|---|---|---|---|---|---|---|---|---|---|---|---|
| MIRACLE-MECFS | f | 30-39 | 21.8 | 24.7 | 22.6 | 21.2 | 20 | 16.9 | 17.9 | 19.9 | 20.2 | 18 | 21.7 | 13.1 | 13 | 13.3 | 11.1 | 15.6 | 11.8 | 15.6 | 16.1 | 18.1 |
| MIRACLE-MECFS | f | 40-49 | 21.8 | 20.2 | 19.6 | 18.6 | 18.3 | 16.7 | 16.5 | 16.1 | 13.3 | 13 | 17.9 | 18.7 | 15.1 | 14.8 | 12.6 | 10.8 | 10.7 | 9.9 | 10.5 | 9.3 |
| MIRACLE-MECFS | f | 60-69 | 21.7 | 20.1 | 19.7 | 18.8 | 18.3 | 18.5 | 16.4 | 17.1 | 16.4 | 15.9 | 19 | 17.2 | 15.6 | 15.4 | 13.1 | 14.9 | 14.1 | 13 | 14.9 | 14.4 |
| MIRACLE-MECFS | f | 50-59 | 21.6 | 24.5 | 17.7 | 14.3 | 16.8 | 11.5 | 13.2 | 15 | 15 | 18.6 | 12.8 | 5.5 | 6.1 | 5.5 | 6.1 | 5.2 | 6.5 | 5.3 | 6 | 6 |
| MIRACLE-MECFS | f | 40-49 | 21.5 | 18.4 | 18.1 | 16.1 | 16.1 | 16.1 | 14.2 | 15.3 | 14.8 | 13.4 | 19.1 | 17.2 | 16.4 | 16.4 | 15.9 | 15.2 | 14 | 14.5 | 14.2 | 13.5 |
| MIRACLE-MECFS | f | 60-69 | 21.3 | 19.5 | 20.5 | 19.4 | 18.3 | 18.5 | 19.2 | 17.2 | 16.6 | 18.6 | 17.2 | 17.8 | 16 | 15.7 | 12 | 11.8 | 14.1 | 12.9 | 14.1 | 13.4 |
| MIRACLE-MECFS | f | 40-49 | 21.2 | 20.6 | 20.9 | 20.6 | 18 | 15.9 | 15.5 | 13.3 | 18 | 16.4 | 13.3 | 12.3 | 9.6 | 9.2 | 8.8 | 8.4 | 8.4 | 7.7 | 8 | 8 |
| MIRACLE-MECFS | f | 30-39 | 20.7 | 21.5 | 18.3 | 18.8 | 16.2 | 16.1 | 15.8 | 15.2 | 15.8 | 14.8 | 22.4 | 19.2 | 19.9 | 19.6 | 19.5 | 19 | 18.3 | 17.5 | 17 | 16.5 |
| MIRACLE-MECFS | f | 50-59 | 20.5 | 24.5 | 21.6 | 21.6 | 19 | 19 | 18.3 | 18 | 18.3 | 18 | 22.1 | 19.1 | 19.8 | 17.1 | 17.7 | 16.3 | 15.8 | 16.5 | 16.2 | 14.9 |
| MIRACLE-MECFS | f | 40-49 | 20.4 | 16.9 | 17.3 | 13.4 | 12.2 | 11 | 10.5 | 9.2 | 10.6 | 8.6 | 8.5 | 6.6 | 6.7 | 4 | 4.9 | 4.7 | 3 | 2.7 | 3.6 | 3.6 |
| MIRACLE-MECFS | f | 50-59 | 20.3 | 14.7 | 11 | 16.8 | 10.8 | 9.3 | 9 | 9.5 | 9.4 | 7 | 15 | 14.2 | 12.7 | 13.5 | 13.1 | 15.4 | 10.5 | 14.2 | 7.8 | 8.4 |
| MIRACLE-MECFS | f | 40-49 | 20.3 | 13.3 | 15.6 | 11.2 | 8.9 | 7.8 | 6.8 | 6.8 | 6.9 | 6.9 | 3.6 | 3.2 | 3 | 3.5 | 5.3 | 3 | 3.5 | 4.2 | 3.3 | 2.7 |
| MIRACLE-MECFS | f | 40-49 | 20 | 22 | 19.2 | 20.1 | 21.6 | 19.3 | 17 | 14.9 | 12.5 | 14.3 | 17.3 | 11.7 | 14.9 | 16.7 | 12.1 | 14.8 | 10.8 | 11.1 | 12.2 | 10.4 |
| MIRACLE-MECFS | f | 50-59 | 19.7 | 12.8 | 12.2 | 12.1 | 10.5 | 10.4 | 8.8 | 7.4 | 7.1 | 8.7 | 22.3 | 12.2 | 11.9 | 8.9 | 13.2 | 11.9 | 11.5 | 12.4 | 8 | 10.6 |
| MIRACLE-MECFS | f | 40-49 | 19.1 | 13.6 | 10.4 | 10.9 | 10.9 | 8.4 | 9.6 | 9.9 | 12 | 11.8 | 7.3 | 6.8 | 5.3 | 4.2 | 4.4 | 4.4 | 4.8 | 4.5 | 4.8 | 4.8 |
| MIRACLE-MECFS | f | 50-59 | 18.3 | 19.8 | 19.5 | 16 | 16.8 | 17.2 | 15.5 | 15.2 | 15.5 | 18.4 | 19 | 15.1 | 15.9 | 14.1 | 11.4 | 14.2 | 12.5 | 14.1 | 14.2 | 11.3 |
| MIRACLE-MECFS | f | 40-49 | 18.1 | 20.2 | 20.6 | 13.6 | 13.9 | 15.7 | 14 | 14.2 | 13.8 | 11.2 | 15 | 12.2 | 14.3 | 12.1 | 12.7 | 11 | 10.3 | 11.6 | 9.8 | 8.8 |
| MIRACLE-MECFS | f | 20-29 | 17.5 | 18.2 | 19.4 | 15.3 | 16.1 | 13.8 | 11.2 | 14.1 | 11.2 | 12.8 | 8.7 | 8.4 | 6.6 | 11 | 6.7 | 8.1 | 6.9 | 8 | 7.7 | 7.9 |
| MIRACLE-MECFS | f | 30-39 | 16.6 | 16.2 | 13.3 | 19.4 | 11.6 | 14.3 | 16.7 | 13.1 | 13.8 | 13.6 | 13.8 | 12.5 | 16.4 | 12.9 | 11.3 | 10.3 | 10.8 | 9.9 | 10.4 | 9.7 |
| MIRACLE-MECFS | f | 50-59 | 16.6 | 17.1 | 16.7 | 15.6 | 13.5 | 14.3 | 14.3 | 10.9 | 13.1 | 11.6 | 12.4 | 13.8 | 12.9 | 12.9 | 11.2 | 12.5 | 10.6 | 12.5 | 11.9 | 11.9 |
| MIRACLE-MECFS | f | 50-59 | 16.5 | 18.9 | 16.5 | 17.5 | 16.4 | 15.8 | 16.4 | 14.6 | 14.9 | 13.9 | 16.2 | 14.5 | 15.6 | 15.8 | 12.8 | 14.3 | 13.2 | 14.2 | 13.4 | 13.6 |
| MIRACLE-MECFS | f | 40-49 | 16.1 | 9.2 | 11.8 | 11.7 | 9.5 | 11.5 | 10.5 | 9.2 | 9.6 | 10.4 | 9.5 | 8.8 | 10.3 | 10.5 | 11.3 | 10.3 | 8 | 11.6 | 11.9 | 11.7 |
| MIRACLE-MECFS | f | 50-59 | 16 | 11.4 | 13.1 | 12.4 | 12.9 | 13.3 | 12.9 | 13.1 | 12.2 | 11.4 | 21.8 | 13.9 | 11.9 | 10.6 | 9.2 | 7.9 | 8.7 | 8.1 | 7 | 8.1 |
| MIRACLE-MECFS | f | 40-49 | 15.6 | 13.7 | 13.5 | 15.5 | 15.4 | 14.9 | 12.6 | 10.5 | 13.2 | 10 | 14.1 | 16.3 | 16 | 14.4 | 13.7 | 14.2 | 13.4 | 13.4 | 13.8 | 12.2 |
| MIRACLE-MECFS | f | 50-59 | 15.5 | 13.8 | 11.4 | 9.2 | 7.7 | 7.2 | 7.8 | 7 | 5.6 | 6.1 | 5.3 | 5.2 | 3.6 | 4.1 | 4 | 3.8 | 3.8 | 3.7 | 3.4 | 3.2 |
| MIRACLE-MECFS | f | 50-59 | 15.1 | 15.1 | 15.6 | 15.1 | 14.2 | 14.7 | 14.1 | 14.3 | 15.3 | 13.5 | 17.4 | 14.5 | 13 | 14.8 | 13.5 | 14.2 | 13 | 12.5 | 12 | 12.2 |
| MIRACLE-MECFS | f | 40-49 | 15 | 11.1 | 16.8 | 13.3 | 9.7 | 6.4 | 7.4 | 8.3 | 9.1 | 7.7 | 8.8 | 9.2 | 6 | 6.2 | 7.9 | 7.4 | 6.7 | 7.8 | 8.5 | 6.6 |
| MIRACLE-MECFS | f | 50-59 | 15 | 20.7 | 17.2 | 19.1 | 17.7 | 16.9 | 16.7 | 15.6 | 17 | 15.8 | 15.3 | 15.9 | 14.9 | 12.1 | 15.2 | 12.3 | 9.7 | 8 | 10.3 | 10.7 |
| MIRACLE-MECFS | f | 50-59 | 14.5 | 12.9 | 8.9 | 9.5 | 8.1 | 7.3 | 7.3 | 6 | 5.4 | 4.7 | 14.8 | 10.4 | 12.6 | 9 | 11.5 | 9 | 8.5 | 8 | 7.2 | 7 |
| MIRACLE-MECFS | m | 30-39 | 14.3 | 15 | 15.6 | 16.1 | 15.3 | 16.5 | 18.6 | 18.7 | 17.7 | 18.7 | 13.1 | 13.7 | 16.7 | 15.1 | 13.3 | 14.1 | 15 | 14.6 | 17.9 | 16.4 |
| MIRACLE-MECFS | f | 30-39 | 14.3 | 17.7 | 17.7 | 13.2 | 12.8 | 14.5 | 12.6 | 11.2 | 11.2 | 12.2 | 12.5 | 10.4 | 9.9 | 10.8 | 11.1 | 11.5 | 9.7 | 10.6 | 10.6 | 11.4 |
| MIRACLE-MECFS | f | 40-49 | 13.7 | 8.3 | 8.4 | 9.8 | 9.3 | 9.8 | 8.6 | 8.4 | 7.4 | 8.3 | 6.8 | 7.3 | 7 | 7 | 7.1 | 6.7 | 6.6 | 6.8 | 5.9 | 6.4 |
| MIRACLE-MECFS | f | 50-59 | 13.1 | 9.3 | 8.9 | 5.6 | 6.9 | 6.9 | 8.5 | 8.1 | 8.7 | 8.4 | 10.8 | 5 | 4.9 | 4.6 | 7.6 | 6.6 | 7.3 | 5.5 | 5.6 | 7.5 |
| MIRACLE-MECFS | f | 60-69 | 13 | 12.1 | 12.2 | 11.2 | 11.4 | 10.7 | 11.5 | 10.1 | 8.7 | 10.3 | 13.5 | 12.8 | 12.2 | 13.2 | 11.7 | 10.8 | 11.2 | 10.9 | 10.5 | 10.4 |
| MIRACLE-MECFS | f | 40-49 | 13 | 15.5 | 13.7 | 14.8 | 15.5 | 8.6 | 8.3 | 8.7 | 6.9 | 7.5 | 6 | 4.6 | 6.6 | 4.2 | 5.8 | 5.2 | 2.7 | 3.4 | 2.7 | 3.1 |
| MIRACLE-MECFS | f | 20-29 | 12.3 | 12 | 10.9 | 10.3 | 10.7 | 11.1 | 9.4 | 8.6 | 8.6 | 10 | 8 | 9 | 8.8 | 8.1 | 8.2 | 8 | 7.6 | 7.9 | 8.3 | 7.3 |
| MIRACLE-MECFS | f | 50-59 | 12 | 13.9 | 10.4 | 9.8 | 10.3 | 10.1 | 8.3 | 9.9 | 6.2 | 5.9 | 7.4 | 7.9 | 7.9 | 8.4 | 6.1 | 4.2 | 4.9 | 5.8 | 5 | 4.8 |
| MIRACLE-MECFS | f | 30-39 | 11.2 | 10.2 | 10.1 | 14.4 | 6.8 | 12 | 11.1 | 12.5 | 12.9 | 12.9 | 7.7 | 7.8 | 5.7 | 5.9 | 8.1 | 5.8 | 7.1 | 6.9 | 7.3 | 6.4 |
| MIRACLE-MECFS | f | 40-49 | 11 | 11.1 | 11.3 | 7.8 | 10.1 | 10.9 | 11 | 7.5 | 5.8 | 5.3 | 14.5 | 5.8 | 2.4 | 6.1 | 5.7 | 7.9 | 9.4 | 5.2 | 5.6 | 6.7 |
| MIRACLE-MECFS | f | 50-59 | 10.7 | 10.7 | 10.6 | 11.2 | 9.9 | 9.4 | 9.6 | 9.4 | 8.1 | 8.5 | 9.9 | 9.3 | 9.9 | 9.7 | 7.5 | 7.7 | 6.3 | 7.9 | 6.9 | 5.8 |
| MIRACLE-MECFS | m | 60-69 | 8.8 | 7.1 | 6.6 | 4.5 | 4.3 | 4.3 | 5.3 | 3.8 | 4.1 | 3.9 | 12.5 | 4.6 | 4.7 | 5.6 | 5.1 | 5.1 | 4.8 | 4.7 | 3.7 | 3.3 |
| MIRACLE-MECFS | f | 50-59 | 8.2 | 5.6 | 4.5 | 5.2 | 5.5 | 4 | 5.1 | 4.9 | 3.1 | 4.8 | 9.3 | 5.8 | 3.8 | 3.2 | 2.8 | 2.3 | 2.6 | 1.9 | 1.8 | 2.1 |
| MIRACLE-MECFS | f | 40-49 | 7.1 | 8 | 6.8 | 6.5 | 6.1 | 6.3 | 6.6 | 6.9 | 6.4 | 10.2 | 6.9 | 6.3 | 5.9 | 5.4 | 4.7 | 4.8 | 4.9 | 5.2 | 4.4 | 4.7 |
| MIRACLE-MECFS | f | 50-59 | 6.9 | 5.5 | 4.9 | 4.3 | 4 | 4.3 | 4.3 | 3.4 | 4.1 | 4.4 | 4 | 3.6 | 3.7 | 4 | 3 | 3.5 | 3.1 | 3.3 | 3.2 | 3 |
| MIRACLE-MECFS | f | 30-39 | 5.8 | 5.4 | 4.7 | 4.6 | 4 | 3.9 | 3.9 | 3.6 | 3.3 | 4.2 | 4.1 | 3.6 | 3.2 | 3.4 | 3.3 | 3.5 | 2.6 | 2.6 | 2.5 | 2.5 |
| MIRACLE-MECFS | f | 60-69 | 3.9 | 5.2 | 5.4 | 4.4 | 4.6 | 3.6 | 4.1 | 4.2 | 4.9 | 5 | 8.9 | 4.9 | 3.5 | 3.7 | 3.3 | 3.8 | 3.6 | 2.7 | 3.1 | 3 |
| MIRACLE-MECFS | f | 40-49 | 3.6 | 4.8 | 4.1 | 5.5 | 3.9 | 3.2 | 3.4 | 3.6 | 3.6 | 3.9 | 3.4 | 3.5 | 3.4 | 3.6 | 3.5 | 3.2 | 3.3 | 3.6 | 3.6 | 3.4 |
| MIRACLE-HC | m | 60-69 | 72.5 | 65.2 | 63.2 | 62.5 | 67.5 | 67.3 | 66.1 | 63.3 | 64.2 | 61.1 | 66.2 | 64.6 | 58.7 | 60.5 | 58.5 | 55.3 | 53.6 | 46.1 | 58.3 | 51.7 |
| MIRACLE-HC | m | 30-39 | 64.6 | 64 | 63.5 | 62.2 | 56.6 | 58.8 | 56.4 | 47.7 | 56.7 | 58.3 | 53.4 | 52 | 43.1 | 48.8 | 50 | 52.1 | 42.4 | 42.7 | 49.1 | 43 |
| MIRACLE-HC | m | 30-39 | 63.7 | 49.8 | 49 | 52.4 | 54.1 | 49.5 | 47.4 | 50.2 | 49.9 | 44 | 59.7 | 54.3 | 46.4 | 50.2 | 49.5 | 42.8 | 42.6 | 39.7 | 46.2 | 41.3 |
| MIRACLE-HC | m | 30-39 | 56.3 | 54.2 | 52 | 47.9 | 48.5 | 49.1 | 47.7 | 47 | 43 | 44.1 | 60.1 | 55.1 | 49.4 | 45.7 | 43.7 | 41 | 41.1 | 42.1 | 39.3 | 38.7 |
| MIRACLE-HC | m | 60-69 | 54.1 | 51 | 50.4 | 49.3 | 46.7 | 45.4 | 45 | 43.6 | 44.1 | 43.7 | 53.9 | 53.5 | 51.7 | 49.5 | 47.8 | 47.2 | 43.5 | 44 | 43 | 42.3 |
| MIRACLE-HC | m | 20-29 | 51.3 | 58.2 | 54.7 | 51.5 | 47.4 | 46.4 | 47.3 | 47.8 | 49.1 | 48.6 | 55.6 | 47.5 | 48.8 | 46.9 | 42.5 | 37.7 | 40.2 | 38.3 | 44.1 | 41.9 |
| MIRACLE-HC | m | 40-49 | 51.3 | 46.7 | 55.6 | 51.4 | 46.5 | 47.5 | 46.1 | 47.9 | 41.4 | 41.9 | 54.8 | 49.3 | 49.5 | 46.5 | 46.6 | 45.3 | 43.8 | 41.3 | 43.7 | 43.6 |
| MIRACLE-HC | m | 20-29 | 50.3 | 49.4 | 50.2 | 49.9 | 48.6 | 49.8 | 51.3 | 47.7 | 49 | 48.4 | 51.8 | 50.3 | 48 | 49.3 | 45.3 | 46.6 | 49.7 | 48.2 | 45.2 | 45.8 |
| MIRACLE-HC | m | 20-29 | 49.8 | 49.2 | 45.8 | 45.6 | 47.2 | 40.3 | 42.2 | 34.8 | 39.7 | 44.1 | 52.1 | 51.9 | 52.7 | 45.5 | 38 | 36.4 | 42.4 | 43.1 | 37 | 35.3 |
| MIRACLE-HC | m | 60-69 | 46.7 | 44.1 | 40.3 | 39.8 | 39.4 | 38.9 | 37.3 | 37.1 | 34.9 | 37.5 | 47.1 | 45.8 | 45.7 | 45 | 42.6 | 41.6 | 39.1 | 38.8 | 33.6 | 33.2 |
| MIRACLE-HC | m | 20-29 | 46.2 | 42.4 | 45.1 | 33.4 | 28.2 | 37.2 | 34.5 | 42.9 | 39.3 | 43.4 | 50.7 | 49 | 44 | 43.8 | 42.8 | 40.3 | 37.3 | 40.4 | 42.4 | 37.8 |
| MIRACLE-HC | m | 20-29 | 45.5 | 53.8 | 51.6 | 48.2 | 41 | 48.4 | 48.3 | 47.8 | 46.8 | 46.4 | 54.2 | 51.7 | 52.4 | 49.5 | 47 | 46 | 46.5 | 47.9 | 47.2 | 47.1 |
| MIRACLE-HC | m | 30-39 | 44 | 44.3 | 43.8 | 42.4 | 42.3 | 38.9 | 38.3 | 39.6 | 39.3 | 38.9 | 43.1 | 43.1 | 40.9 | 40.8 | 35.8 | 39.3 | 35 | 31 | 33.5 | 34.9 |

| | | | | | | | | | | | | | | | | | | | | | | |
|---|---|---|---|---|---|---|---|---|---|---|---|---|---|---|---|---|---|---|---|---|---|---|
| MIRACLE-HC | m | 60-69 | 43.1 | 48 | 51.9 | 54.6 | 53 | 45.6 | 46.7 | 46.7 | 39.6 | 49.2 | 42.1 | 54 | 46.9 | 36.3 | 41.6 | 39.1 | 49.5 | 44.7 | 45.8 | 36.6 |
| MIRACLE-HC | m | 40-49 | 43.1 | 39.8 | 42.6 | 42.4 | 43.3 | 39.5 | 44.2 | 39.8 | 41 | 44.4 | 42.5 | 42.2 | 41.7 | 44.7 | 44.4 | 42.3 | 44.7 | 41.3 | 43.9 | 46.9 |
| MIRACLE-HC | m | 20-29 | 41.7 | 52.3 | 52.3 | 54.1 | 53.7 | 52.1 | 51.1 | 48.8 | 50.7 | 45.8 | 54.9 | 53.6 | 54.3 | 48.4 | 50.5 | 50.2 | 51 | 51 | 48.6 | 47.1 |
| MIRACLE-HC | m | 30-39 | 40.7 | 37.5 | 31.4 | 32.5 | 31.8 | 30 | 29.1 | 29.1 | 29.5 | 26.3 | 35.7 | 32.8 | 34.4 | 29.8 | 24.1 | 29.1 | 27.5 | 26.8 | 24.4 | 25.1 |
| MIRACLE-HC | f | 20-29 | 40.5 | 38.9 | 39.8 | 38.5 | 39.1 | 35.8 | 39.8 | 36.9 | 33.9 | 39.5 | 43.9 | 39.4 | 40.2 | 37.3 | 37.9 | 38.1 | 35.5 | 38.9 | 39.7 | 39.4 |
| MIRACLE-HC | f | 20-29 | 40.5 | 40.4 | 37.9 | 36.1 | 35.6 | 33.7 | 33.7 | 30.8 | 30.8 | 34 | 40.5 | 38.3 | 35.9 | 34.8 | 34.5 | 33.6 | 33.4 | 31.6 | 31.2 | 31.9 |
| MIRACLE-HC | f | 30-39 | 40.1 | 41.4 | 42.9 | 43.1 | 42.2 | 40.7 | 41.6 | 40.6 | 38.5 | 40.3 | 44 | 44.8 | 44.7 | 42.1 | 42.1 | 41.3 | 41.5 | 39.4 | 41 | 38.4 |
| MIRACLE-HC | m | 30-39 | 39.7 | 37.2 | 38.4 | 34.3 | 27.4 | 36.9 | 32.8 | 34.5 | 33.9 | 33.5 | 48.3 | 45.8 | 39.3 | 42.8 | 36.7 | 39 | 39.1 | 35.2 | 34.5 | 38.7 |
| MIRACLE-HC | f | 30-39 | 39.4 | 36.2 | 34.3 | 33.3 | 32.1 | 30.4 | 29.6 | 29.1 | 28.6 | 28.2 | 36.4 | 35.5 | 35.8 | 33 | 29.7 | 27.9 | 28.9 | 28.6 | 28.7 | 28.3 |
| MIRACLE-HC | m | 20-29 | 38.3 | 36 | 29.2 | 32.8 | 35.8 | 34.6 | 34.3 | 34.2 | 31.1 | 32.4 | 40.9 | 39.4 | 38.2 | 34.8 | 35.1 | 31.7 | 30.5 | 29.2 | 34.4 | 31.3 |
| MIRACLE-HC | f | 50-59 | 38.1 | 31.4 | 37.1 | 34.2 | 35.7 | 35.9 | 35.5 | 32.9 | 33.2 | 33.1 | 36 | 35.9 | 34.4 | 33.1 | 30.1 | 32 | 31.6 | 28.8 | 30.4 | 28.5 |
| MIRACLE-HC | f | 20-29 | 36.9 | 35.6 | 33.2 | 32.6 | 31.5 | 27.9 | 31.3 | 30.3 | 28.1 | 28.8 | 29.8 | 31.9 | 31.3 | 28.3 | 28.2 | 27.4 | 26.4 | 26 | 26.5 | 24.9 |
| MIRACLE-HC | m | 40-49 | 36 | 34.2 | 33.1 | 36.3 | 29.6 | 30.3 | 27.2 | 25.3 | 33.2 | 34.7 | 36.7 | 37.7 | 35.1 | 33.2 | 29 | 32 | 31.5 | 29.2 | 30.6 | 31.1 |
| MIRACLE-HC | f | 30-39 | 35.5 | 33 | 32.4 | 29.9 | 29.7 | 28.4 | 28.7 | 29 | 28.1 | 28.1 | 32.5 | 33.5 | 31.9 | 30.2 | 28.8 | 28.3 | 28.1 | 27.1 | 25.6 | 26.9 |
| MIRACLE-HC | f | 30-39 | 34.8 | 30.4 | 30.4 | 28.5 | 28.1 | 23.5 | 29.9 | 29.3 | 32.9 | 29.7 | 33.3 | 31.1 | 33.5 | 32.1 | 30.9 | 31 | 28.5 | 27.9 | 27.1 | 25.5 |
| MIRACLE-HC | f | 20-29 | 34.7 | 31.8 | 31.3 | 30.7 | 30.3 | 28.6 | 27.6 | 27 | 27 | 23.9 | 28.9 | 31 | 27.9 | 28.5 | 27.5 | 25.9 | 27.4 | 27.6 | 26.9 | 27 |
| MIRACLE-HC | m | 20-29 | 34.5 | 36.3 | 35.1 | 44.5 | 37.1 | 38.9 | 42.4 | 39.8 | 37.1 | 41.9 | 42.7 | 36.6 | 40.7 | 41.9 | 44.9 | 35.6 | 34.3 | 45.7 | 44.6 | 43.1 |
| MIRACLE-HC | f | 20-29 | 33.9 | 30.2 | 32.8 | 34.8 | 37.1 | 34.6 | 34.1 | 29 | 33.1 | 29.9 | 32.2 | 34.5 | 36.8 | 35.4 | 33.9 | 32.8 | 33.5 | 33.9 | 31.6 | 27.4 |
| MIRACLE-HC | m | 40-49 | 33.7 | 36.5 | 35.5 | 35.2 | 37.2 | 36.5 | 34.7 | 33.7 | 33.2 | 35.6 | 43.2 | 38.7 | 33.5 | 34.7 | 37.2 | 34.5 | 37.9 | 33.6 | 33.5 | 33.7 |
| MIRACLE-HC | f | 50-59 | 33.1 | 31.5 | 31.3 | 29.8 | 28.7 | 29.7 | 28 | 27.9 | 25.7 | 28 | 21.4 | 23.1 | 25.5 | 24.8 | 23.6 | 24.6 | 24.2 | 23.7 | 25.9 | 25.4 |
| MIRACLE-HC | f | 50-59 | 32.7 | 38.5 | 35.9 | 38.1 | 33.9 | 34.9 | 35.7 | 32.8 | 33.2 | 31.5 | 30.3 | 33.3 | 30.9 | 35 | 32.1 | 31.7 | 32.2 | 30.6 | 28.9 | 27.6 |
| MIRACLE-HC | f | 20-29 | 32.1 | 28.9 | 26 | 25.1 | 22.9 | 22.9 | 22.8 | 23.6 | 23.8 | 23.9 | 28.3 | 28.1 | 25 | 23.5 | 23.2 | 21.1 | 23.4 | 22.1 | 21.2 | 21.9 |
| MIRACLE-HC | f | 60-69 | 32.1 | 29 | 30.2 | 29.1 | 27 | 28.7 | 27.9 | 24.6 | 25.4 | 25.6 | 29.9 | 32.2 | 29.6 | 27.6 | 27.7 | 27.2 | 27.9 | 24.8 | 25.6 | 27.8 |
| MIRACLE-HC | f | 20-29 | 31.9 | 31.4 | 32 | 30.6 | 30.1 | 30.3 | 29.6 | 28.6 | 28 | 28.3 | 30.3 | 27.8 | 29.4 | 28.3 | 27.9 | 28 | 27 | 25.9 | 26.5 | 25.8 |
| MIRACLE-HC | f | 40-49 | 31.6 | 29.1 | 27.5 | 25.6 | 23.5 | 22.4 | 22.4 | 21 | 20 | 20.7 | 27.5 | 24.8 | 24.8 | 21.1 | 22.2 | 21.8 | 20.7 | 18.9 | 17.6 | 19 |
| MIRACLE-HC | f | 50-59 | 31.2 | 29.5 | 24.4 | 25.8 | 24.8 | 24.5 | 22.7 | 22.3 | 23.8 | 23.9 | 27.9 | 27.4 | 26.4 | 24.7 | 23.5 | 23 | 21.9 | 20.2 | 21.6 | 22.8 |
| MIRACLE-HC | m | 50-59 | 31 | 26 | 27.8 | 26.4 | 27 | 26 | 27.6 | 26.2 | 27.3 | 26.7 | 24.8 | 30.5 | 28.9 | 26.3 | 27.8 | 26.8 | 28 | 26 | 25.9 | 26.4 |
| MIRACLE-HC | f | 20-29 | 30.7 | 33.1 | 35.6 | 31.6 | 30.4 | 30.7 | 29.3 | 33.8 | 31.6 | 30.2 | 36.2 | 35.5 | 35.2 | 32.5 | 31 | 30.8 | 31.1 | 29.1 | 29.9 | 28.6 |
| MIRACLE-HC | f | 50-59 | 30.6 | 28.1 | 28.7 | 25.9 | 27 | 25.9 | 26.4 | 24.1 | 25.6 | 28.5 | 34.7 | 31.7 | 31.1 | 28.2 | 27 | 27.6 | 27.1 | 30.2 | 27.1 | 27.2 |
| MIRACLE-HC | f | 20-29 | 30.3 | 30.7 | 29.9 | 27.9 | 26.7 | 25.9 | 25.1 | 25.4 | 24.6 | 23.6 | 28.6 | 26.2 | 26.3 | 23.7 | 23 | 21.4 | 21.3 | 18.8 | 16.9 | 18.6 |
| MIRACLE-HC | f | 60-69 | 29.6 | 27.6 | 26.4 | 25.9 | 24.3 | 24.3 | 24.4 | 22.2 | 25.9 | 26.1 | 25.1 | 24.7 | 23.5 | 23.1 | 21.7 | 22 | 21.4 | 20.8 | 20.5 | 20.4 |
| MIRACLE-HC | f | 20-29 | 29.5 | 30.1 | 30 | 29.3 | 30.5 | 22.8 | 23.8 | 25.6 | 22.9 | 21.8 | 30.8 | 28.6 | 28.5 | 29.6 | 25.9 | 28.4 | 26.9 | 28.2 | 26 | 25.2 |
| MIRACLE-HC | f | 40-49 | 29.3 | 25.4 | 28.4 | 23.4 | 24.2 | 23.7 | 23.6 | 24.6 | 24.9 | 24.4 | 30 | 29.1 | 28.9 | 26.3 | 24.7 | 23.9 | 26.3 | 23.6 | 23.4 | 21.2 |
| MIRACLE-HC | f | 20-29 | 29.3 | 29.9 | 28.6 | 31.2 | 29.5 | 29.1 | 29.1 | 29.2 | 28.1 | 28 | 28.7 | 27.2 | 27 | 24.9 | 25.2 | 25.2 | 25.2 | 25.2 | 26.1 | 25.3 |
| MIRACLE-HC | f | 60-69 | 29.2 | 23.8 | 22.7 | 21.8 | 20 | 18.5 | 21.2 | 18.7 | 22.7 | 24.6 | 29.5 | 26.4 | 27.2 | 23.6 | 23.7 | 22.5 | 22.3 | 21.6 | 20.8 | 25.9 |
| MIRACLE-HC | f | 20-29 | 28.8 | 29.2 | 28.9 | 25.5 | 25 | 24.1 | 24.7 | 23.7 | 22.6 | 22.9 | 29.2 | 32.7 | 28.8 | 28.2 | 25.3 | 25.7 | 23.5 | 22.3 | 19.9 | 20.5 |
| MIRACLE-HC | f | 40-49 | 28.3 | 28.2 | 27.8 | 27.3 | 27 | 26.4 | 27.2 | 27.2 | 27 | 24.8 | 31.6 | 30.3 | 28.4 | 26.7 | 25.5 | 25.5 | 24.9 | 24.6 | 23.5 | 24.3 |
| MIRACLE-HC | f | 20-29 | 28.2 | 26.7 | 25.5 | 27.9 | 22.8 | 22.2 | 22.1 | 21.6 | 23.1 | 22.2 | 22.9 | 20.9 | 20 | 19.7 | 21.1 | 19 | 23.2 | 24.1 | 20.7 | 22.8 |
| MIRACLE-HC | f | 30-39 | 28.1 | 27.7 | 28.9 | 28.6 | 26.8 | 25.1 | 26.2 | 26.5 | 26.1 | 25.7 | 30 | 30 | 27.5 | 27.7 | 24.8 | 24.1 | 27.1 | 24.8 | 24.8 | 24.1 |
| MIRACLE-HC | f | 30-39 | 28 | 26.6 | 23 | 22.8 | 23.6 | 24.9 | 21.1 | 24.9 | 22.4 | 22.9 | 28.7 | 26.3 | 26.6 | 24.5 | 23.4 | 23.2 | 22 | 22 | 20.3 | 25.3 |
| MIRACLE-HC | f | 30-39 | 27.8 | 25.6 | 22.9 | 21.9 | 20.6 | 19 | 19 | 17.9 | 18.2 | 16.8 | 24.7 | 21.3 | 21.7 | 22 | 21 | 19.3 | 20.1 | 20.1 | 19.1 | 18.3 |
| MIRACLE-HC | f | 20-29 | 27.7 | 28.1 | 27.4 | 24.5 | 27.1 | 24.7 | 24.8 | 24.9 | 23.6 | 23.6 | 27.3 | 24.9 | 24.9 | 26.6 | 24.8 | 23.8 | 23 | 26.6 | 21.6 | 23.1 |
| MIRACLE-HC | f | 40-49 | 27 | 28.9 | 26.7 | 30.2 | 29.9 | 29.5 | 33.3 | 32.9 | 29.8 | 30.5 | 35.7 | 31.5 | 31 | 33.8 | 33.2 | 29.5 | 30.1 | 30.8 | 31.8 | 31.1 |
| MIRACLE-HC | f | 40-49 | 26.8 | 28.4 | 26.4 | 29 | 28.1 | 26.8 | 28.1 | 27.6 | 26.9 | 27.6 | 28.7 | 29.7 | 26.9 | 27.5 | 28.8 | 25.7 | 27.5 | 26.5 | 27 | 26.2 |
| MIRACLE-HC | f | 60-69 | 26.6 | 26.2 | 24.6 | 25 | 23.7 | 22.6 | 22.1 | 23 | 22.4 | 20.4 | 22.3 | 20.8 | 18.6 | 19.7 | 18.6 | 17 | 17.4 | 15.9 | 16.4 | 15.9 |
| MIRACLE-HC | f | 20-29 | 26.5 | 24.6 | 24 | 24.7 | 23.8 | 21.6 | 22.5 | 22 | 22.4 | 22.6 | 22.9 | 23.3 | 21.8 | 19 | 19.2 | 20.7 | 18.7 | 18.1 | 18.7 | 20.9 |
| MIRACLE-HC | f | 20-29 | 26.1 | 22.6 | 23.6 | 23.4 | 23.3 | 22.5 | 21.7 | 21.7 | 20.3 | 21.9 | 26.8 | 24.6 | 23.7 | 23.9 | 22.9 | 21.9 | 22 | 21.4 | 20.7 | 20.4 |
| MIRACLE-HC | f | 60-69 | 25.8 | 24.8 | 24.3 | 24.6 | 24 | 24 | 24.5 | 23.6 | 24.3 | 25.1 | 25.7 | 24.7 | 25.1 | 25.2 | 25 | 24.5 | 24.9 | 24.5 | 25.3 | 24.6 |
| MIRACLE-HC | f | 20-29 | 25.7 | 25.2 | 22 | 21.7 | 22.2 | 20.2 | 19.5 | 20.9 | 23.7 | 24.2 | 30.5 | 24 | 20.4 | 18.6 | 17.6 | 19.6 | 17.5 | 18.6 | 14.1 | 13 |
| MIRACLE-HC | f | 20-29 | 25.1 | 25.4 | 27.3 | 26.4 | 26.2 | 25.9 | 26.4 | 26.1 | 27.7 | 27 | 29.9 | 25.5 | 24.7 | 27.2 | 26.2 | 23.2 | 24.4 | 25.1 | 24.6 | 27.6 |
| MIRACLE-HC | f | 20-29 | 25 | 22 | 21.6 | 18.7 | 21.6 | 21.6 | 20.3 | 20.9 | 19.4 | 20.7 | 24.9 | 24.2 | 24.7 | 22.3 | 20.7 | 18.8 | 22 | 18.6 | 18.4 | 19.2 |
| MIRACLE-HC | f | 60-69 | 24.9 | 25.2 | 24.6 | 23.7 | 23.7 | 22.7 | 21.9 | 21.4 | 20.9 | 20.4 | 24.8 | 23.8 | 22.4 | 21.8 | 21.5 | 23.5 | 21.7 | 19.7 | 21.6 | 20 |
| MIRACLE-HC | f | 30-39 | 24.4 | 25.8 | 23 | 21.3 | 22.3 | 20.2 | 21.4 | 21.6 | 21.2 | 21.7 | 27.2 | 24.6 | 22.2 | 23.3 | 19.9 | 25.1 | 24.9 | 23.7 | 23.6 | 22.6 |
| MIRACLE-HC | f | 20-29 | 23.6 | 24.5 | 26.8 | 27.4 | 22.1 | 25.5 | 23.5 | 20.8 | 22.9 | 19.2 | 26.5 | 26 | 27.4 | 25.3 | 22.9 | 25.8 | 25.8 | 24.2 | 23 | 20.4 |
| MIRACLE-HC | f | 50-59 | 23.3 | 23.2 | 22.9 | 21.7 | 22.6 | 24 | 22 | 22.1 | 20.4 | 20.9 | 27.3 | 26.8 | 27.2 | 24.8 | 23.6 | 22.9 | 20.6 | 19.1 | 23.4 | 19.6 |
| MIRACLE-HC | f | 40-49 | 21.7 | 19.9 | 21.7 | 20.6 | 18 | 20.6 | 19.2 | 19.9 | 19.9 | 20.5 | 25 | 24.4 | 22.3 | 23.1 | 21.1 | 20.4 | 21.4 | 21.4 | 20.7 | 20.5 |
| MIRACLE-HC | m | 20-29 | 21.6 | 20.2 | 20.4 | 20.5 | 17.7 | 25.3 | 22.6 | 21.5 | 22.6 | 22.9 | 22.3 | 23.6 | 27.2 | 25 | 24.6 | 23.4 | 21.5 | 21.2 | 21.9 | 20.9 |
| MIRACLE-HC | f | 10-19 | 20.9 | 21.3 | 21.7 | 18.2 | 18.5 | 17.1 | 17.1 | 17.6 | 16.3 | 18.8 | 25.9 | 23.8 | 23 | 21.1 | 21.1 | 20.6 | 18.8 | 18.2 | 17.7 | 21 |
| MIRACLE-HC | f | 50-59 | 18.7 | 19.8 | 15.3 | 16.3 | 17.7 | 17.8 | 18.1 | 16.3 | 14.8 | 15.9 | 16.3 | 14.9 | 14.3 | 15.6 | 13.5 | 14.9 | 15.4 | 14.2 | 15.2 | 12.9 |
| MIRACLE-HC | f | 70-79 | 17.6 | 17.5 | 17.5 | 15.4 | 17.2 | 16.4 | 16.3 | 16.3 | 16.9 | 16.4 | 17.1 | 18.2 | 16.2 | 17.1 | 16.8 | 15.4 | 17.7 | 18.7 | 19.7 | 20.7 |

| | | | | | | | | | | | | | | | | | | | | | | |
|---|---|---|---|---|---|---|---|---|---|---|---|---|---|---|---|---|---|---|---|---|---|---|
| MIRACLE-HC | f | 20-29 | 15.7 | 16.8 | 16.7 | 15.9 | 13.8 | 14.1 | 16.1 | 15.1 | 15.7 | 19.5 | 18.5 | 18.9 | 20.2 | 16.6 | 16.1 | 17 | 16.5 | 17.1 | 16.9 | 20.2 |
| MIRACLE-HC | f | 50-59 | 15.4 | 18.5 | 16.9 | 19.3 | 17.2 | 18.7 | 18.2 | 20.4 | 18.6 | 20.7 | 20.3 | 19.1 | 19.3 | 23.2 | 21.6 | 22.3 | 23 | 20.8 | 21.7 | 22.8 |
| MIRACLE-HC | m | 60-69 | 14.3 | 22.2 | 21.6 | 19.9 | 23.9 | 23 | 26.9 | 28.3 | 30.4 | 31.1 | 19.8 | 22.6 | 25.7 | 28.2 | 27.5 | 28.2 | 27.5 | 31.4 | 34.2 | 35 |
| HC-submax | f | 30-39 | 13.1 | 12 | 12.2 | 6.7 | 7.1 | 9.3 | 9.7 | 10 | 7.7 | 10.9 | 7.8 | 7.5 | 5.9 | 5 | 5.2 | 5.8 | 8.8 | 6.8 | 5.9 | 6.5 |
| HC-submax | f | 30-39 | 9.1 | 7.6 | 9.7 | 5.4 | 6.5 | 6 | 6 | 5.2 | 5.1 | 7.6 | 6.4 | 9.4 | 7 | 5.1 | 6.4 | 4.8 | 5.3 | 5.1 | 6 | 6.5 |
| HC-submax | m | 60-69 | 19.7 | 22.4 | 18.4 | 17.1 | 18.5 | 16.7 | 15.8 | 13.3 | 11.9 | 14.7 | 15.9 | 18.7 | 19.8 | 17 | 16.6 | 18.1 | 15 | 15.3 | 15 | 14.5 |
| HC-submax | m | 50-59 | 19.5 | 18.1 | 22.5 | 15.4 | 16.8 | 12.9 | 10.8 | 12.7 | 10.3 | 11.9 | 15.8 | 17.6 | 11.7 | 10.6 | 11.6 | 10.9 | 11 | 11.3 | 11.9 | 11.8 |
| HC-submax | f | 30-39 | 10.1 | 10.7 | 6.9 | 6.9 | 7.5 | 6.8 | 7.3 | 7.8 | 9 | 11.1 | 8.7 | 8.6 | 8.3 | 10.5 | 8.7 | 6.3 | 7.3 | 7.2 | 6.3 | 6.2 |
| HC-submax | m | 30-39 | 11.7 | 11.4 | 12.5 | 12.7 | 15.2 | 16.1 | 16.6 | 12.4 | 16.5 | 16.7 | 13 | 19.4 | 20.3 | 20.4 | 19.7 | 18.6 | 21.4 | 22.5 | 19.7 | 19.4 |
| HC-submax | f | 20-29 | 9.1 | 10.4 | 12.4 | 10.1 | 12.2 | 12.9 | 10.6 | 8.3 | 9.2 | 7.7 | 7 | 7.2 | 6.2 | 5.5 | 6.3 | 6.7 | 5.7 | 7.8 | 8.7 | 8.5 |
| HC-submax | f | 20-29 | 9.3 | 13.4 | 15.1 | 13.4 | 11.5 | 11.1 | 11.4 | 9.9 | 9.9 | 9.1 | 12.4 | 10.2 | 12.1 | 9 | 7.6 | 9.6 | 8.3 | 12.7 | 11.9 | 12.3 |
| HC-submax | m | 20-29 | 16.4 | 15.2 | 13.7 | 13.5 | 14.1 | 13 | 14.2 | 9.6 | 10.2 | 10.4 | 14.3 | 17.6 | 12.4 | 12.3 | 11.5 | 11.7 | 11.4 | 11.7 | 11.6 | 11.1 |
| HC-submax | f | 30-39 | 1.3 | 1.2 | 2.4 | 4.3 | 6.2 | 6 | 4 | 4.7 | 4.7 | 4.5 | 2.7 | 3.6 | 4.1 | 3.1 | 3.2 | 3.2 | 3.1 | 3.6 | 3.7 | 3.4 |
| HC-submax | m | 40-49 | 37.5 | 37 | 42.3 | 35.9 | 33.6 | 33.9 | 35.3 | 32.4 | 34 | 32.7 | 41.2 | 37.3 | 41.9 | 35.2 | 33.1 | 34.2 | 35.2 | 33.8 | 30.8 | 33.1 |
| HC-submax | m | 30-39 | 25 | 40.4 | 39.9 | 36.5 | 35.7 | 37.9 | 29.5 | 34.8 | 31.2 | 27.3 | 30.2 | 42.1 | 32.5 | 35.5 | 31.7 | 29.5 | 36.2 | 39.9 | 43.3 | 33.9 |
| HC-submax | f | 40-49 | 10.6 | 13 | 16.8 | 16.3 | 14.6 | 14.5 | 14.7 | 16.4 | 16.8 | 16.5 | 12 | 11.2 | 8.4 | 8.1 | 7.6 | 8.9 | 7.5 | 9 | 8.3 | 7.8 |
| HC-submax | f | 50-59 | 10.8 | 10.6 | 10 | 9 | 8.1 | 8.1 | 7.8 | 8.6 | 8.1 | 7.2 | 8.6 | 8.2 | 7 | 5.9 | 7.4 | 4.8 | 3.7 | 3.7 | 4.4 | 4.2 |
| HC-submax | m | 20-29 | 14.7 | 14.7 | 15.2 | 18.5 | 15.9 | 15.5 | 14.3 | 16.4 | 15.1 | 15.3 | 28.1 | 19.2 | 13 | 12.8 | 12 | 14 | 12.8 | 14 | 13.4 | 15.1 |
| HC-submax | m | 20-29 | 14 | 15.2 | 15.7 | 13 | 10 | 9.7 | 10.8 | 13.4 | 14.1 | 13.4 | 13.4 | 13.4 | 9.9 | 10.5 | 10.4 | 9.4 | 11.4 | 11.9 | 11 | 11.4 |
| HC-submax | f | 30-39 | 7.7 | 12.4 | 9.4 | 13.5 | 12.8 | 10.7 | 10.9 | 8.9 | 8.3 | 13.8 | 16.5 | 12.6 | 13.7 | 11.5 | 11.5 | 12.3 | 12 | 9.7 | 9 | 10.7 |
| HC-submax | f | 20-29 | 6.6 | 5.9 | 6.8 | 5.1 | 6.1 | 6.5 | 6.7 | 6.2 | 6.6 | 6.1 | 4.5 | 5 | 5.6 | 4.9 | 5.8 | 5.8 | 5.1 | 5.6 | 5.6 | 6.3 |
| HC-submax | f | 20-29 | 6.2 | 3.6 | 2.8 | 2.2 | 1.9 | 1.9 | 3 | 2.2 | 2.5 | 2.1 | 2.7 | 1.2 | 1.1 | 2 | 2.1 | 1.9 | 1.4 | 1.6 | 1.3 | 1.5 |
| HC-submax | m | 20-29 | 20.3 | 20.1 | 20.9 | 21.1 | 19.6 | 20.8 | 19.9 | 20.9 | 17.9 | 18.6 | 29.8 | 24.1 | 21.4 | 18.5 | 18.3 | 16.5 | 16.4 | 13.8 | 15.6 | 14.1 |
| HC-submax | m | 20-29 | 5.8 | 6.8 | 6.1 | 5.5 | 4.7 | 5.4 | 5.1 | 5.6 | 4.7 | 5 | 10.3 | 7.5 | 6.6 | 7.5 | 7.3 | 7.3 | 6.5 | 6.8 | 6.8 | 8 |
| HC-submax | m | 40-49 | 4.6 | 6.8 | 8.3 | 8.5 | 8.9 | 8 | 8 | 9.7 | 9.2 | 9.1 | 6.3 | 6.3 | 6.5 | 6.2 | 6 | 7.6 | 5.9 | 7.1 | 7.8 | 7.7 |
| HC-submax | m | 50-59 | 21.3 | 19.4 | 20.2 | 19.6 | 18.3 | 19.6 | 20.1 | 19.7 | 20.6 | 18.1 | 17.8 | 15 | 18.8 | 18.9 | 21.6 | 22.2 | 21.9 | 18.3 | 19 | 21.6 |
| HC-submax | f | 20-29 | 9.2 | 14.8 | 15.1 | 16.1 | 17.1 | 20.3 | 21.9 | 20.7 | 16.2 | 16.5 | 9.3 | 10.9 | 9.4 | 11.4 | 10.1 | 9.5 | 9.5 | 10.2 | 10.7 | 10.9 |
| HC-submax | m | 20-29 | 14.2 | 12.4 | 12.5 | 12.7 | 10.2 | 10.3 | 9.7 | 10.1 | 11.2 | 12.1 | 22 | 16.9 | 16.2 | 15.9 | 12.9 | 12.9 | 16.2 | 14.4 | 15.9 | 15.6 |
| HC-submax | f | 20-29 | 14.2 | 16.2 | 14.2 | 10 | 10.1 | 7.5 | 8.2 | 10.1 | 7.7 | 7.3 | 8.6 | 5.7 | 7.8 | 4.6 | 5 | 3.8 | 4.7 | 3.1 | 4.2 | 4.1 |
| HC-submax | f | 20-29 | 9.1 | 9.8 | 8.8 | 8.5 | 8.6 | 6 | 5.1 | 5.8 | 5.4 | 4 | 8.7 | 6.3 | 4.2 | 3.9 | 4 | 5.1 | 2.7 | 2.7 | 3 | 3.3 |
| HC-submax | f | 20-29 | 17.6 | 18.7 | 17.8 | 16.3 | 17.3 | 16.8 | 13.9 | 13.1 | 15.3 | 15.2 | 17.8 | 14.5 | 15.2 | 16.1 | 12.9 | 10.7 | 13.6 | 12.6 | 13.2 | 13.1 |
